# A class of deep intronic *IGHMBP2* variants activate a shared cryptic splice donor, enabling correction of select variants with a single antisense oligonucleotide

**DOI:** 10.64898/2026.04.20.26351111

**Authors:** Sarah Silverstein, Andrew D Nguyen, Rotem Orbach, Sandra Donkervoort, Thomas Cassini, Mary Koziura, Véronique Bolduc, Audrey M Winkelsas, Ester Masati, Shreya Nandi, George Harmison, Brian Johnson, Kory Johnson, Sarah E Kargbo-Hill, Jason J. Bussgang, Jahan Misra, Ishaan Sharma, Jordan E Bontrager, David N. Herrmann, Francesco Vetrini, Erin Conboy, Adam Comer, Kayla Treat, Katelyn Payne, Khurram Liaqat, Aneesh G. Patankar, Alayne P Meyer, Daniel C. Koboldt, Anne M. Connolly, Richard Shell, Anthony R Miller, Pimchanok Kulsirichawaroj, Oranee Sanmaneechai, Kullasate Sakpichaisakul, Kyeyoon Park, Yan Li, Diana Bharucha-Goebel, William Macken, Anna Sarkozy, James Polke, Adnan Y Manzur, A. Reghan Foley, Katherine R Chao, Sarah Neuhaus, David R Adams, Michael Ward, Carsten G. Bönnemann, Christopher Grunseich

**Affiliations:** Neuromuscular and Neurogenetic Disorders of Childhood Section, NINDS, NIH, Bethesda MD; Rutgers New Jersey Medical School, Newark, NJ; Undiagnosed Diseases Program, NIH, Bethesda MD; Inherited Neuromuscular Disease Unit, Neurogenetics Branch, NINDS, NIH Bethesda MD; Department of Pediatrics, Vanderbilt University Medical Center, Nashville TN; Bioinformatics Core, NINDS, NIH Bethesda MD; University of Rochester Medical Center, Department of Neurology, NY; Undiagnosed Rare Disease Clinic (URDC), Department of Medical and Molecular Genetics, Indiana University School of Medicine, Indianapolis, Indiana 46202, USA; Department of Neurology, Indiana University School of Medicine, Indianapolis, Indiana 46202, USA; Division of Genetic and Genomic Medicine, Nationwide Children’s Hospital, Columbus, OH; Center for Gene Therapy, Abigail Wexner Research Institute at Nationwide Children’s Hospital, Columbus, OH; The Steve and Cindy Rasmussen Institute for Genomic Medicine, Abigail Wexner Research Institute, Nationwide Children’s Hospital, 700 Children’s Drive, Columbus, OH 43205, USA; Department of Pediatrics, The Ohio State University College of Medicine, Columbus, OH; Division of Neurology, Nationwide Children’s Hospital, Columbus, OH; Division of Pulmonary, Sleep and CF, Nationwide Children’s Hospital, Columbus OH; Department of Pediatrics, Faculty of Medicine Siriraj Hospital, Mahidol University, Bangkok, Thailand; Siriraj Center of Research Excellence in Neuromuscular Disease, Faculty of Medicine Siriraj Hospital, Mahidol University, Bangkok, Thailand; Pediatric Precision Medicine Center, Department of Pediatrics, Faculty of Medicine, Siriraj Hospital, Mahidol University, Bangkok, Thailand; Department of Pediatrics, Queen Sirikit National Institute of Child Health, Ministry of Public Health, Bangkok, Thailand; College of Medicine, Rangsit University, Bangkok, Thailand; Stem Cell Unit, NINDS, NIH Bethesda MD; Proteomics Core Facility, NINDS, NIH Bethesda MD; Department of Neuromuscular diseases, UCL Queen Square Institute of Neurology, Queen Square House, London WC1N 3BG, UK; Dubowitz Neuromuscular Centre, Great Ormond Street Hospital, Institute of Child Health, University College London, London, UK; National Hospital for Neurology and Neurosurgery and North Thames Genomics Laboratory Hub Rare Disease Laboratory, Queen Square, London, UK; Broad Institute of MIT/Harvard, Boston MA; Office of the clinical director, NHGRI, NIH Bethesda MD

**Keywords:** Rare Diseases, RNA-Seq, Diagnosis, Neuromuscular Diseases, iPSC derived Motor Neurons, Splicing, Precision Medicine, Noncoding variants, Long-Read Sequencing

## Abstract

Biallelic disease-causing variants in *IGHMBP2* cause spinal muscular atrophy with respiratory distress type I (SMARD1) and Charcot-Marie-Tooth type 2S (CMT2S). We present 12 unrelated patients with clinically suspected *IGHMBP2*-related-disease, each carrying a variant deep in intron 8 of *IGHMBP2* (c.1235+1076G>A (n=6), c.1235+450G>A (n=5), and c.1235+894C>A (n=1)), along with a known deleterious variant in trans. To assess aberrant pathogenic splicing induced by these deep intronic variants in a relevant model, patient-derived induced pluripotent stem cells were differentiated into motor neurons (iMNs). Long-read RNA sequencing revealed introduction of different pseudoexons by each variant: c.1235+450G>A (626bp), c.1235+1076G>A (112bp and 77bp) and c.1235+894C>A (182bp). Although each variant utilizes a unique splice acceptor site, they all activate the same cryptic donor site, enabling a therapeutic approach to redirect aberrant splicing for all the variants using a single shared antisense oligonucleotide (ASO). Treatment of iMNs with this single ASO restored full-length IGHMBP2 protein in c.1235+894G>A and c.1235+1076G>A by decreasing the use of the novel acceptor site. In contrast, ASO treatment did not correct the splicing in c.1235+450G>A, suggesting that additional splice correction will be needed for this specific variant. A CRISPR interference screen of IGHMBP2 loss-of-function in iMNs identified ribonucleoprotein complex biogenesis (RNP), and rRNA and tRNA processing as top pathways implicated in motor neuron vulnerability. Proteomics and transcriptomics analysis of successfully treated patient iMNs revealed correction of RNP biogenesis and rRNA processing defects. This study highlights the importance of characterizing deep intronic variants in disease-relevant cells to assist the diagnostic process and inform therapeutics development.

**One Sentence Summary:** Intron 8 of *IGHMBP2* is a hotspot for splice activating pathogenic variants causing SMARD1 and CMT2S, which can be targeted with a single antisense oligonucleotide to correct the aberrant splicing, increase protein and restore cellular function in patient derived motor neurons.

## Introduction

Biallelic pathogenic variants in *IGHMBP2* result in two clinically distinct but related diseases. Pathogenic variants in *IGHMBP2* were initially associated in 2001 with spinal muscular atrophy with respiratory distress type 1 (SMARD1), an early-onset disorder characterized by severe weakness and respiratory failure, now accounting for approximately 71% of reported *IGHMBP2*-related cases (*1–5*). In contrast, Charcot–Marie–Tooth disease type 2S (CMT2S), associated with *IGHMBP2* in 2014, presents later in childhood with a predominantly motor axonal neuropathy and classically no respiratory involvement (*6*). CMT2S has been associated with variants predicted to preserve higher levels of IGHMBP2 protein; however, emerging cases in which identical variants result in divergent clinical presentations challenge a strict genotype–phenotype correlation, suggesting that additional modifiers may be at play (*6–9*). Although gene replacement and splice-correction approaches have shown efficacy in preclinical models, and clinical trials are currently underway, disease-modifying therapies are not yet available for this group of disorders (*10–16*).

Despite its established disease association, the specific function of IGHMBP2 in motor neurons remains incompletely understood (*11*). IGHMBP2 is a ubiquitously expressed, ATP-dependent RNA/DNA helicase belonging to the UPF1-related helicase superfamily, yet loss-of-function (LOF) disproportionately affects alpha motor neurons (*11, 17*). Proposed roles span multiple aspects of RNA biology, including transcription, RNA processing, RNA decay, and translation, with the strongest evidence supporting a role in translational regulation through ribosomal RNA (rRNA) processing and direct ribosome association (*11, 17–24*). Most studies localize IGHMBP2 to the cytoplasm, despite proposed roles in rRNA and tRNA processing that require nuclear or nucleolar localization. However, its localization and function have not been well characterized in motor neurons, the disease-relevant cell type (*17–20*). The development of high-throughput CRISPR interference (CRISPRi) screens and their application in i^3^Neurons (integrated, inducible, and isogenic iPSC-derived neurons) offers a powerful tool to investigate IGHMBP2 pathways that are essential to different neuronal subtypes (*25–27*). This approach could shed light on the unique vulnerability of motor neurons to *IGHMBP2* LOF.

In this study, we describe a region deep within intron 8 of *IGHMBP2* in which pathogenic splice-active variants were clustered in twelve unrelated patients with compatible clinical phenotypes. Using patient-derived induced lower motor neurons, we evaluated the splicing consequences of these variants using long-read transcriptome sequencing and compared these findings with short-read approaches. We utilized a high-throughput CRISPRi screen to investigate genetic and molecular pathways that may underlie differences in survival between motor and cortical neurons in control and *IGHMBP2* LOF models. The detailed characterization of aberrant splice isoforms caused by intronic variants informed the development of an expanded splice-corrective therapeutic strategy, potentially addressing all variants in this cluster with a single ASO. Correction of *IGHMBP2* splicing in patient-derived iMNs also provides insight into the rescue of vulnerable molecular pathways in a disease-relevant cellular context.

## Results

### Case Series

Exome and/or genome sequencing identified 12 unrelated individuals with rare heterozygous variants of uncertain significance deep in intron 8 of *IGHMBP2* (NM_002180.3) in trans with a pathogenic or likely pathogenic variant (Fig 1A, Table S1, Supplemental Case Reports). All patients presented with symptoms of CMT2S or SMARD1 and electrophysiological findings consistent with a diagnosis of *IGHMBP2-*related-disease (Fig 1A, Table S1). The first variant c.1235+450G>A, was found in five patients (gnomAD total v4.1.0 allele frequency (AF) 0.00008854, 104/985276 alleles, 0 homozygotes reported), the second variant c.1235+894C>A was found in one patient who has been previously reported (*10, 28*) (not present in gnomAD), and the third variant, c.1235+1076G>A was found in six patients (gnomAD v4.1.0 total AF 0.00007098, 15/1289560 alleles, 0 homozygotes reported). Splice prediction scores with SpliceAI (*29*) and Pangolin (*30, 31*) are low, with the strongest prediction of aberrant splicing for the c.1235+1076G>A variant (Table 1). Most patients were diagnosed with CMT2S (75%, n = 9), while those with SMARD1 (25%, n = 3) exhibited delayed respiratory involvement, with onset at ages 3, 4 and 10 years (Table S1).

**Figure 1:**
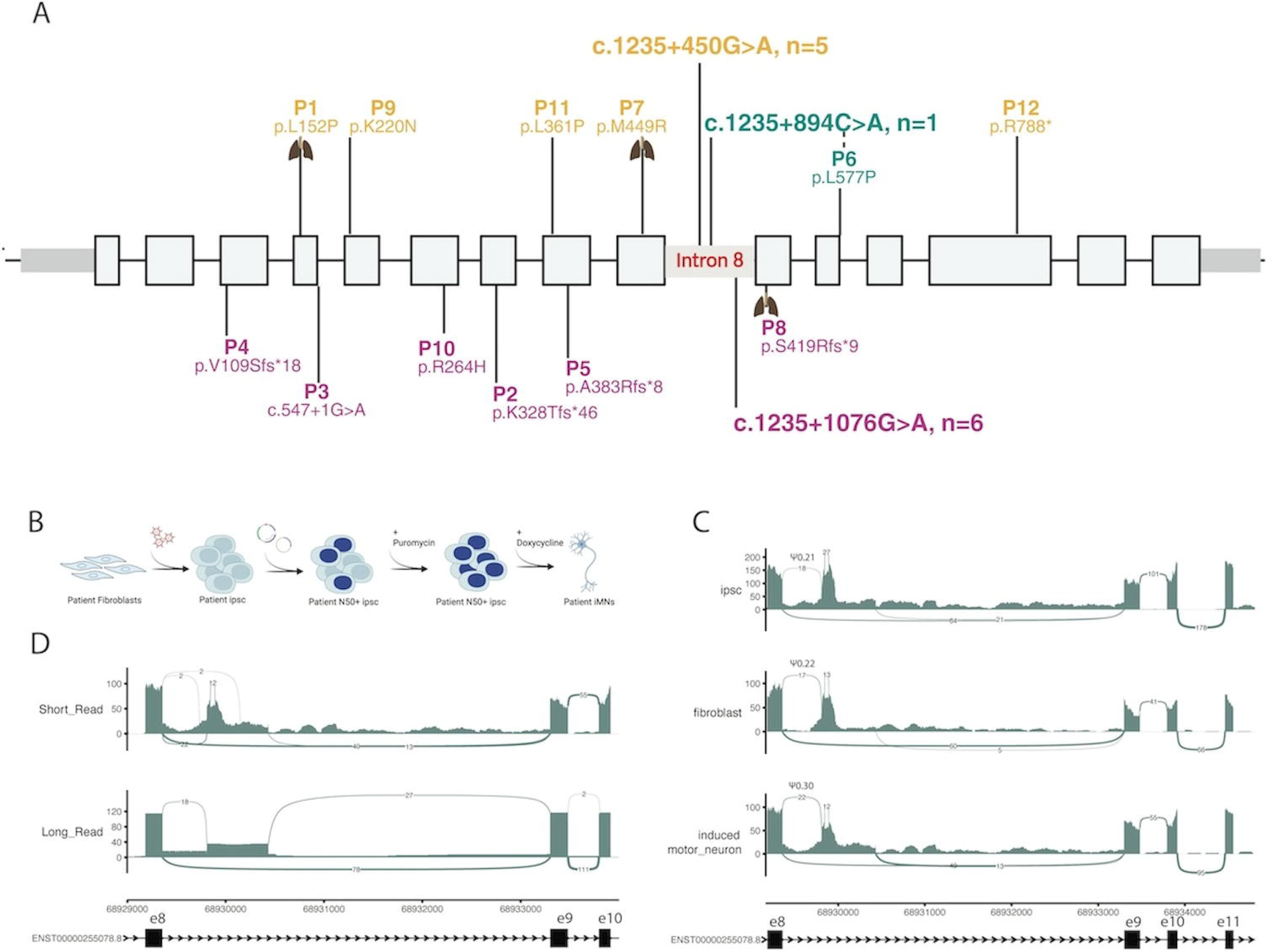
Genetic landscape of patients with IGHMBP2 intron 8 variants and rationale for cell modeling and sequencing selection. A) twelve unrelated patients were identified harboring one pathogenic/likely pathogenic variant in *IGHMBP2* in trans with a variant of uncertain significance deep in intron 8. Patients are colored by deep intron 8 variant, yellow for c.1235+450G>A, teal for c.1235+894C>A and pink for c.1235+1076G>A. Lung icon denotes patients with respiratory involvement. Figure created with Biorender. B) representative patient fibroblasts were collected for each intronic variant, engineered into iPSCs and then differentiated into motor neurons (iMN) with doxycycline. Figure created with Biorender. C) RNA extracted from c.1235+450G>A patient fibroblasts, iPSCs and iMNs were sequenced with short-reads, revealing creation of a novel acceptor site. Percent spliced in (PSI) for each tissue source demonstrates highest use of novel acceptor in iMNs. PSI values are reported for each tissue above the splice junction. D) long read sequencing of RNA from iMNs elucidates a 626bp pseudoexon created due to the c.1235+450G>A variant not detected by short-read sequencing.

**Table 1:**
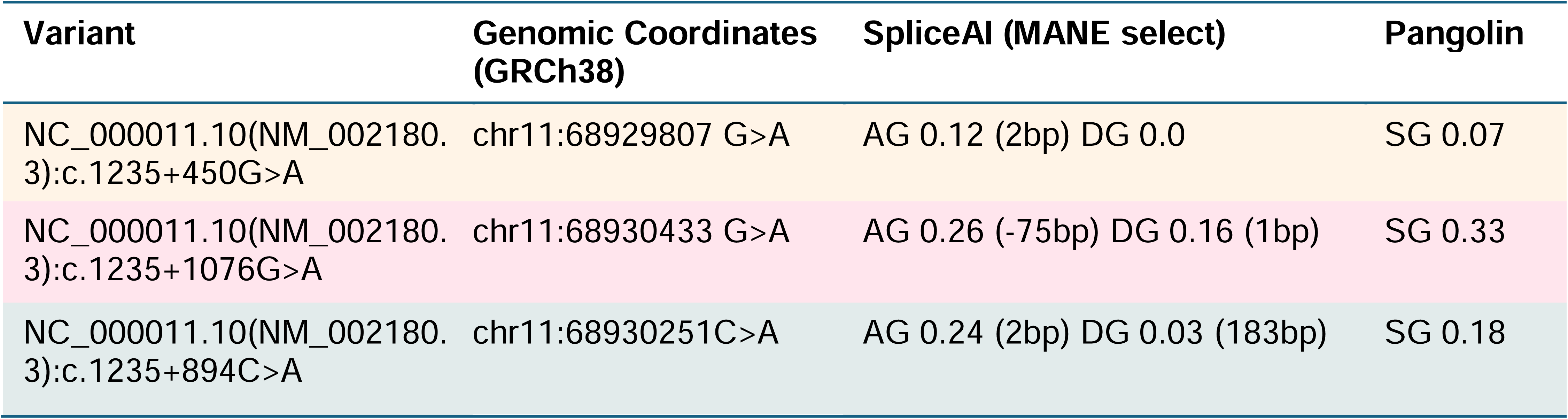
Splice prediction of deep intronic variants in IGHMBP2. AG = acceptor gain, DG =donor gain, SG = splice gain.

### Variant Validation

To assess the functional impact of these variants, three cell models were compared to identify the optimal system for the analysis. Patient-derived fibroblasts carrying the c.1235+450G>A variant (P1) were reprogrammed into iPSCs and differentiated into lower motor neurons (iMNs) using the I^3^LMN inducible system (Fig. 1B) (*27*). Short-read RNA-seq from all three tissue sources showed that the c.1235+450G>A variant generates a novel acceptor, with percent splice-in (PSI) varying by cell type. Splicing was highest in iMNs (PSI 30%) compared with other tissues (PSI 20%; Fig. 1C, Table S2). Although the novel acceptor site was detected by short-read RNA-seq, the full impact of the aberrant splice was obscured by intron retention and multiple nearby splice events (Fig. 1C). Long-read RNA-seq in iMNs provided greater clarity and revealed the boundaries of a 626 bp pseudoexon insertion activated by the c.1235+450G>A variant (Fig. 1D). Based on these findings, variant characterization for the additional variants proceeded using patient-derived iMN models and long-read RNA-seq. Patient-derived iMN models were generated from a representative patient for each of the three variants. Karyotyping confirmed genomic stability (Fig. S1A), and cells were positive for the motor neuron markers HB9 and β-III tubulin (Fig. S1B).

Targeted long-read RNA-seq was used to increase coverage over *IGHMBP2* (Table S3). Reads were haplotype-phased to determine allele-specific splicing contributions from the intron 8 variants (Fig. S2A-C), and allele-specific PSI (asPSI) values were calculated for each cell line. The c.1235+450G>A variant produced a 626 bp pseudoexon (asPSI 23%; Fig. 2A-B, Table S2), while the c.1235+894C>A variant generated a 182 bp pseudoexon (asPSI 49.5%; Fig. 2A-B). These pseudoexons were absent in control samples. The c.1235+1076G>A variant created two overlapping pseudoexons (77 bp, asPSI 26%; 112 bp, asPSI 27.5%) present at very low levels in other patient samples and controls (Fig. 2A-B). All pseudoexons contain premature termination codons when spliced into the normal *IGHMBP2* reading frame and are subject to nonsense-mediated decay. Despite using different acceptor sites, all three variant-created pseudoexons share the same cryptic donor (Fig. 2A-B). Predicted splice sites from SpliceAI were either incorrect (c.1235+450G>A) or only partially correct (c.1235+1076G>A, only the 77 bp pseudoexon was correctly predicted; Table 1, Fig. 2A-B). Notably, a fraction of reads from each variant retained canonical splicing (asPSI 30–76%; Fig. 2B), likely contributing a variable amount of full-length IGHMBP2 protein from this allele.

**Figure 2:**
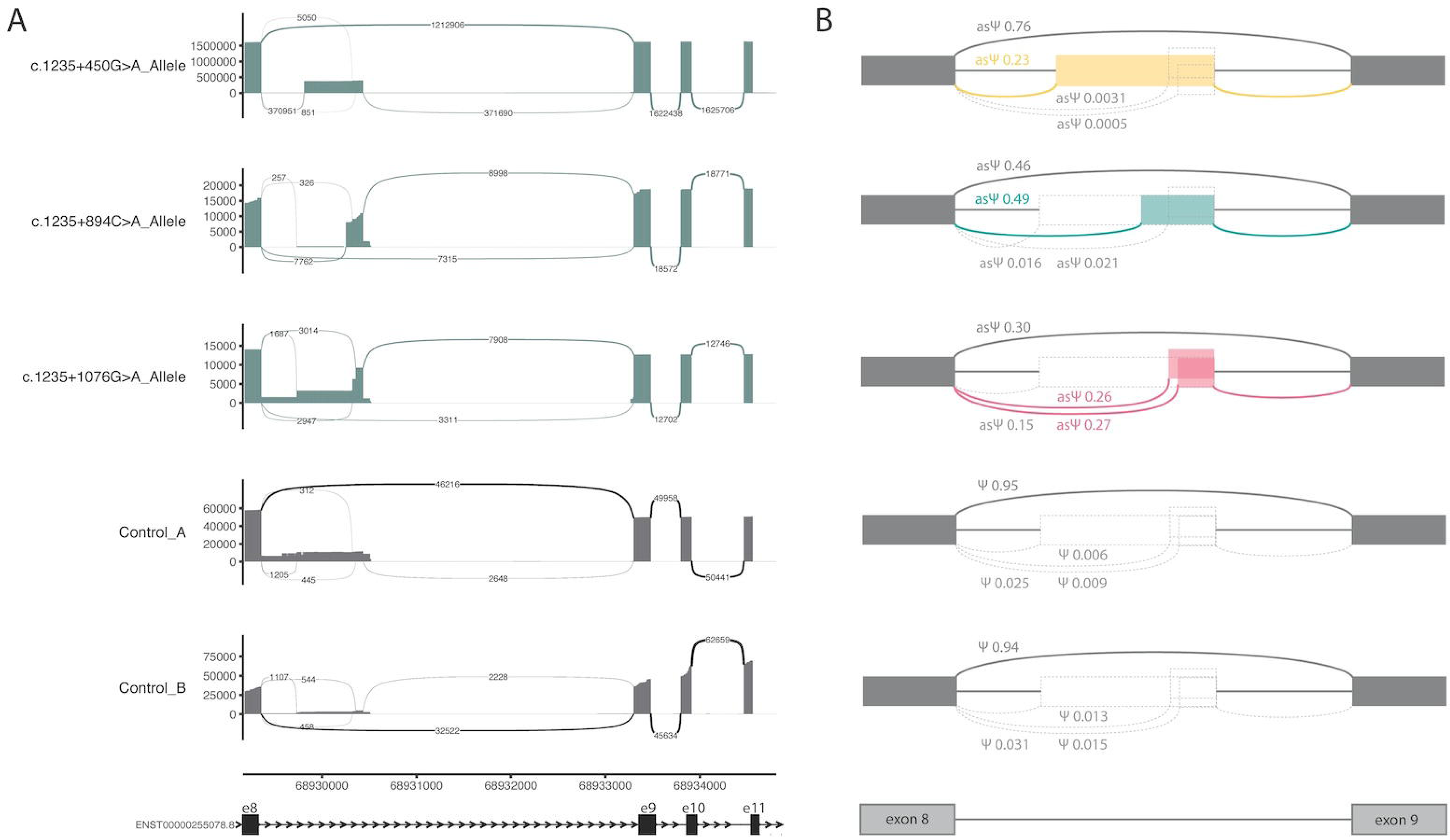
Targeted long read RNAseq from patient derived iMNs highlights aberrant splicing in intron 8 of IGHMBP2. A) sashimi plots from phased long read data demonstrating splicing occurring on intron 8 variant containing allele. Control samples are not phased due to lack of coding variants to perform phasing. B) splice diagrams for each sample, corresponding to the sashimi plot in the same row. Grey splice junctions represent canonical splicing, with corresponding allele-specific PSI (asPSI) values above. Dotted grey boxes and lines demonstrate non-canonical splice events observed at very low levels, with asPSI values in light grey by corresponding dotted splice junctions. Yellow boxes and lines refer to aberrant splicing unique to c.1235+450G>A variants, teal for c.1235+894C>A and pink for c.1235+1076G>A, with corresponding asPSI values in the same colors.

### A single antisense oligonucleotide restores IGHMBP2 protein

Given that all three intronic variants activated the same cryptic donor, an antisense oligonucleotide (ASO) was designed to target a sequence within the shared region (Fig. 3A). Initial testing of the ASO, synthesized as a morpholino (I2-PMO), in c.1235+1076G>A fibroblasts showed a dose-dependent reduction in the 112-bp pseudoexon inclusion compared with a non-targeting control ASO (STD-PMO) at doses ≥200 nM (p < 0.0001; Fig. S3A-B). Following this positive dose response in c.1235+1076G>A fibroblasts, the ASO was synthesized as a Vivo-Morpholino (I2-vPMO), the efficacy of which was then tested in patient-derived iMNs for all three variants, using a single 1μM dose for 96 hours (Fig. 3B).

**Figure 3:**
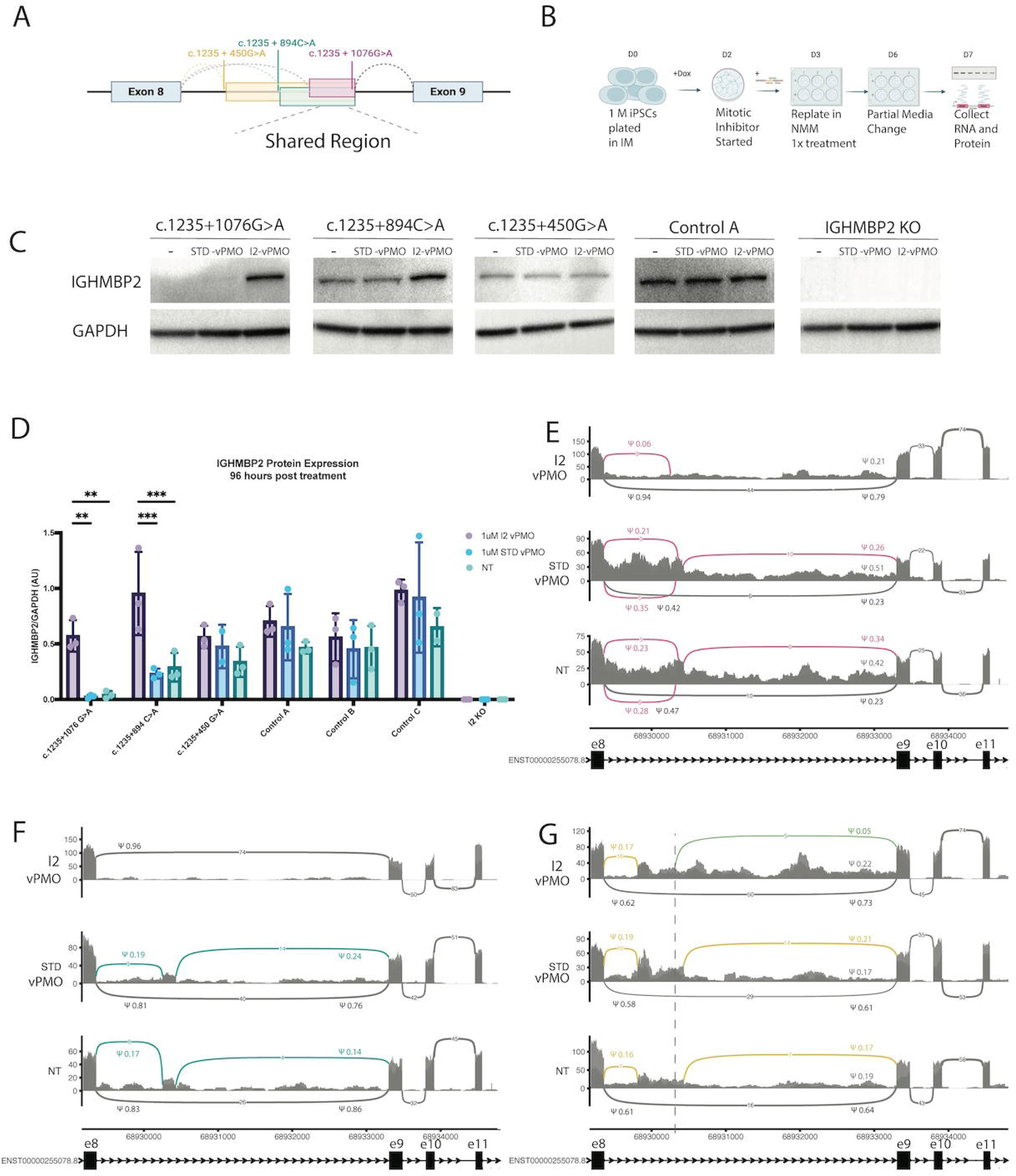
Antisense oligonucleotide treatment of patient-derived iMNs reveals target engagement for two out of three variants. A) Schematic of pseudoexons created by *IGHMBP2* intron 8 variants highlighting the shared cryptic donor and the overlapping region where the IGHMBP2 ASO (I2-vPMO) was designed. Figure created with Biorender. B) Treatment protocol of patient and control iMNs with 1uM I2-vPMO, standard control ASO (STD-vPMO) or no treatment (NT). Cells were treated once on day 3 of differentiation. Treatment experiments were performed in triplicate for each cell line and condition, and both RNA and protein were collected on day 7 post differentiation. Figure created with Biorender. C) Protein analyses via western blot confirmed target engagement for c.1235+894C>A and c.1235+1076G>A variants but not for c.1235+450G>A. D) Western blot bands were quantified and normalized to GAPDH. Each dot represents the average of two technical westerns, and each condition included three biological replicates. Statistical analysis confirmed increase in full-length IGHMBP2 protein for c.1235+1076G>A with I2-vPMO treatment compared to STD-vPMO (two-way ANOVA with Tukey’s multiple comparisons test, df = 40, p=0.0041) and compared to NT ((wo-way ANOVA with Tukey’s multiple comparisons test, df = 40, p=0.0054) and for c.1235+894C>A variants with I2-vPMO treatment compared to STD-vPMO (two-way ANOVA with Tukey’s multiple comparisons test, df = 40, p = 0.0002). and compared to NT (two-way ANOVA with Tukey’s multiple comparisons test, df = 40, p = 0.0005). E,F) Target engagement corrects IGHMBP2 splicing in c.1235+1076G>A (E) and c.1235+894C>A (F) cell lines. I2-vPMO- treated samples showed increased canonical splicing of exons 8-9 compared with STD-vPMO and NT. Each sashimi graph is the aggregate of three replicates, with median splice junction values reported. PSI values were quantified using MAJIQ. Pink and teal splice junctions = aberrant splice events, black splice junctions = canonical splice events. G) Target engagement doesn’t correct *IGHMBP2* splicing in c.1235+450G>A cell line. I2-vPMO-treated samples unchanged usage of novel acceptor in I2-vPMO compared with control treatment. Loss of the cryptic donor is observed in the I2-vPMO condition, with increased intron retention (0.22). Activation of an alternative cryptic donor proximal to the I2-vPMO target is observed (green reads, 5%, dotted line demonstrating distance from targeted cryptic donor). Each graph is the aggregate of three replicates, with median splice junction values reported. PSI values were quantified using MAJIQ. Yellow splice junctions = aberrant splice events. Black splice junctions = canonical splice events.

Target engagement was first assessed at the protein level. *IGHMBP2* knockout cells confirmed antibody specificity (Fig. 3C). Western blot analyses showed statistically significant increased IGHMBP2 protein following I2-vPMO treatment for the c.1235+894C>A (p < 0.0001) and c.1235+1076G>A variants (p < 0.001) (Fig. 3C-D). No treatment effect was observed for the c.1235+450G>A line. To investigate the mechanism underlying increased protein levels, short-read RNAseq was performed. Gene counts for *IGHMBP2* were quantified, normalized and compared between conditions. Normalized *IGHMBP2* gene counts were unchanged in I2-vPMO samples compared with a standard control (STD-vPMO) or non-treated (NT) controls (Fig. S4).

To confirm that increased protein resulted from splice correction, short-read RNA-seq was custom-mapped to known aberrant splice sites and PSI values in intron 8 quantified with MAJIQ. For the c.1235+1076G>A variant, canonical splicing between exon 8 and exon 9 was restored to 94%, compared with 42% and 47% in STD-vPMO and NT samples, respectively (Fig. 3E, Fig. S5A-B). For the c.1235+894C>A variant, canonical splicing was restored to 96% from 81% and 83% in STD-vPMO and NT, respectively (Fig. 3F, Fig. S5C). For the c.1235+450G>A variant, loss of splicing at the shared cryptic donor was observed, indicating expected target engagement (Fig. 3G). Usage of the novel acceptor however remained largely unchanged (17%, 19%, and 16% in I2-vPMO, STD-vPMO, and NT, respectively; Fig. 3G). Additionally, 5% usage of an alternative cryptic donor was detected, along with a 5% increase in distal intron retention (22% in I2-vPMO vs. 17% in STD-vPMO, Fig. 3G). Thus, while the cryptic donor was effectively targeted, the 626-bp pseudoexon was not fully blocked, allowing splicing at the novel acceptor to persist (Fig. 3G).

### CRISPRi Screen identifies cell type-specific vulnerabilities in iMNs

To characterize the cell type-specific consequence from *IGHMBP2* LOF and thereby better assess the consequences of the transcript-directed treatment, we conducted a CRISPRi screen in iPSC-derived cortical and motor neurons (Fig. S6A-B). Control iPSCs expressing NIL (motor neuron) or NGN2 (cortical neuron) were transfected with a transposon vector encoding an enzymatically inactive Cas9 (dCas9) fused to a Krüppel-associated box (KRAB) repressor domain (dCas9-KRAB), enabling targeted transcriptional repression in a single guide RNA (sgRNA)-dependent manner (Fig S6C). This CRISPR interference (CRISPRi) system allows for precise and tunable knockdown of gene expression in iMNs and induced Cortical Neurons (iCNs). Knockdown of gene expression in iCNs via sgRNAs has previously been demonstrated (*27*). The two engineered cell lines allowed for differentiation to iMN and iCN and expression of the dCas9-KRAB construct to facilitate CRISPRi-mediated gene repression. A CRISPRi screen was performed by transducing both iMNs and iCNs with a control sgRNA and a pooled sgRNA library (Figure S6D). Control sgRNA treatment was completed to facilitate comparison with *IGHMBP2* knockdown. To assess the impact of gene knockdown, sgRNA abundance was quantified using next generation sequencing at day 0 (D0), D3, and D14 post differentiation. Reduced abundance of sgRNAs in the D3 or D14 versus D0 samples indicates a negative impact on neuronal viability during differentiation, whereas enrichment at D3 or D14 versus D0 indicates a potential survival advantage from sgRNA-mediated knockdown.

Data analysis was performed using the MAGeCKFlute pipeline to calculate phenotype scores from sgRNA sequence counts (*27*). Redundant but unique sgRNAs targeting the same gene were aggregated and compared against non-targeting negative control sgRNAs to generate phenotype scores and p-values (Fig. 4A). This approach allowed for the identification of genes and pathways essential for iMN and iCN survival. Gene Ontology (GO) analysis was then used to categorize the top 200 genes whose phenotype scores were most affected by the CRISPRi screen in each neuronal subtype and their associated biological processes. GO analysis for iCNs revealed that the top-ranked processes are involved in mRNA processing, tRNA metabolism and cholesterol biosynthesis, aligning with previously published datasets (Fig. 4B) (*27*). GO analysis in iMNs identified RNA splicing, rRNA and tRNA processing among the top pathways implicated by the CRISPRi screen for motor neuron survival (Fig. 4C). A comparative analysis of phenotype scores between iMNs and iCNs was performed to compare the genes and pathways that impact each neuronal subtype. This analysis revealed that a majority of sgRNAs had a negative phenotype score in both iMNs and iCNs at both time points (Fig. S7).

**Figure 4:**
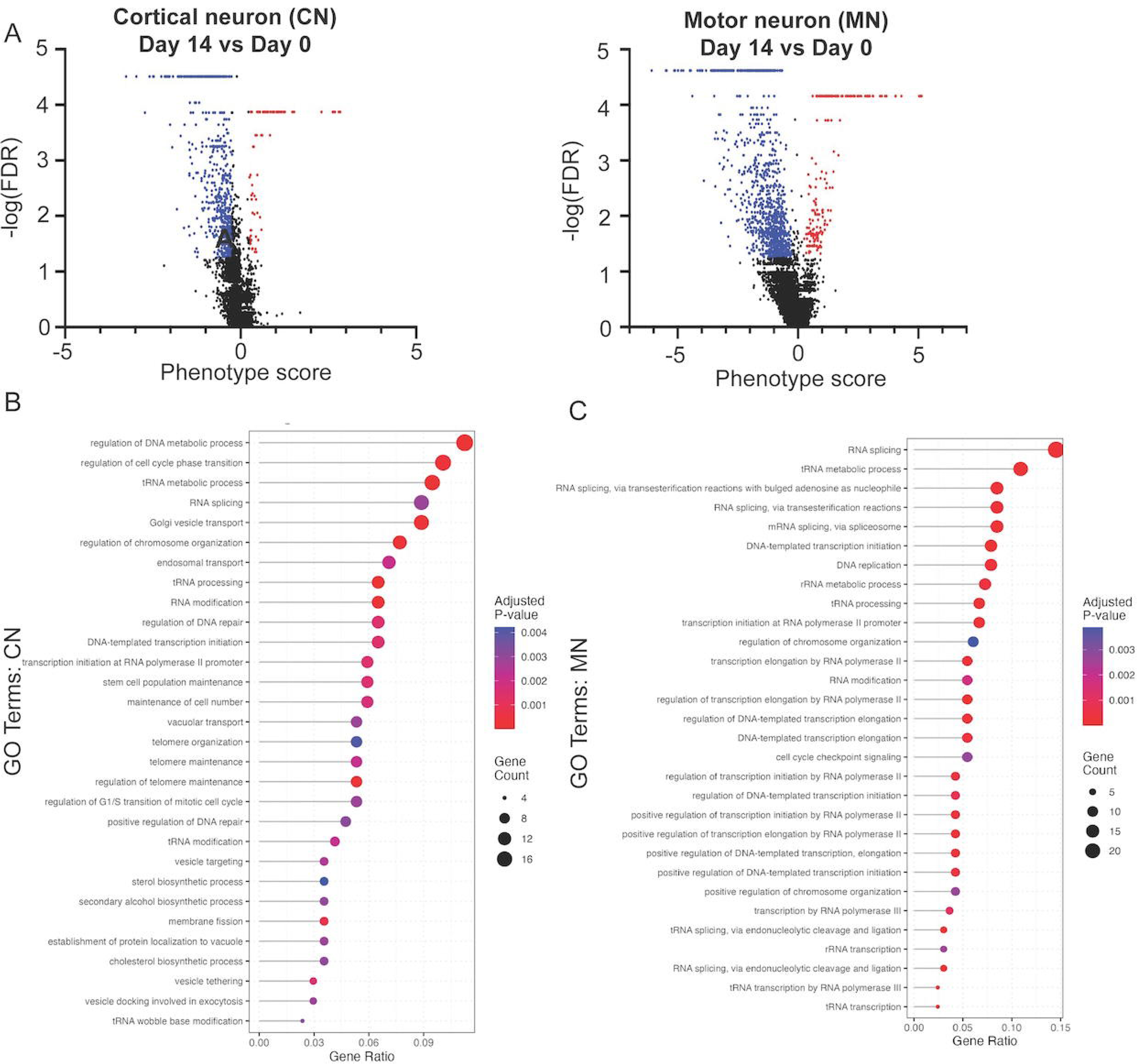
CRISPRi screen identifies pathways selectively vulnerable in iCNs vs iMNs. A) Phenotype scores shows that sgRNA mediated gene knockdown results in decreased survival advantage to cortical and motor neurons. Target genes with -log(FDR)>1.25 and phenotype score <-0.25 indicated in blue and genes with -log(FDR)>1.25 and phenotype score >0.25 indicated in red. Top 200 genes (absolute value of the phenotype score) were annotated for gene ontology pathways, demonstrating unique pathways vulnerable in iCNs versus iMNs. B) For iCNs, the top 30 categories include tRNA processing, RNA splicing, cholesterol biosynthesis. C) For iMNs, top 30 categories include tRNA processing, RNA splicing, and rRNA processing.

### IGHMBP2 knockdown CRISPRi Screen

To investigate how *IGHMBP2* LOF specifically causes motor neuron disease, undifferentiated iPSCs expressing dCas9-KRAB were transduced with a lentiviral vector expressing an sgRNA targeting *IGHMBP2* to generate *IGHMBP2*-knockdown (*IGHMBP2*-KD) iCN and iMN lines (Table S4). *IGHMBP2*-targeting sgRNAs reduced levels of IGHMBP2 protein in undifferentiated iPSCs and differentiated neurons (Fig. S6E). These cells were subsequently transduced with the same pooled sgRNA library used in the control CRISPRi screen, generating double-knockdown lines. Phenotype scores were then computed following the same analysis pipeline as in the control sgRNA neuron CRISPRi screen (Fig. 5A, Table S5)

**Figure 5:**
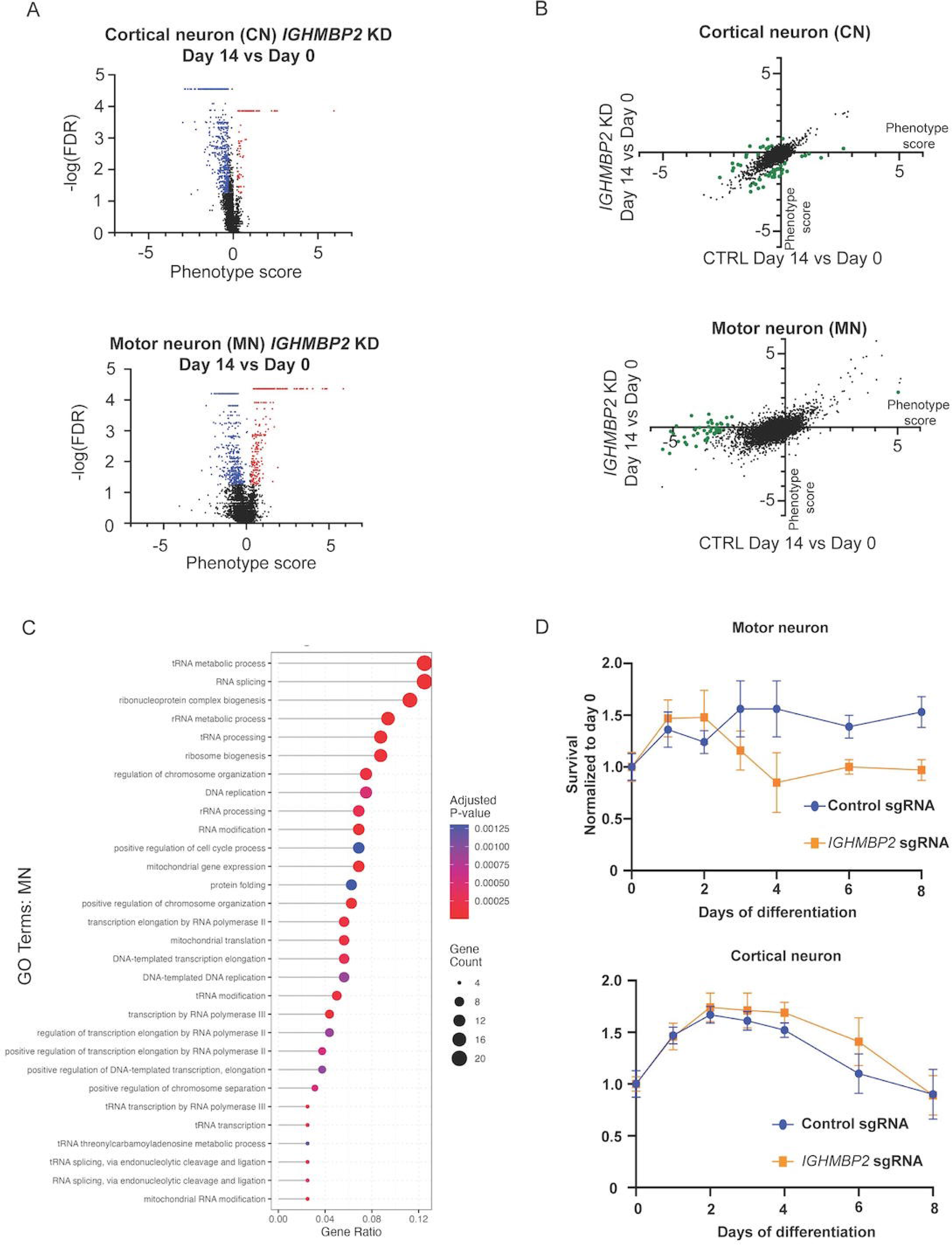
Dual IGHMBP2 knockdown and CRISPRi screen. A) sgRNA phenotype scores for both iCN and iMN’s with *IGHMBP2* KD. B) comparison of phenotype scores to control iMN/ iCN shows mitigation of phenotype scores with *IGHMBP2* KD but not for iCN. Each gene depicted as a separate dot and the top 50 genes most impacted by *IGHMBP2* KD shown enlarged in green. C) gene ontology annotation of mitigated genes in iMN *IGHMBP2*-KD reveal tRNA processing, rRNA processing, and RNP biogenesis among the top 30 pathways. D) survival analysis of iMN and iCN reveal a decrease in survival with *IGHMBP2*-KD for iMNs but not iCNs over 8 days of differentiation. Significant difference is found at day 4 post differentiation (two-way ANOVA with sidak’s multiple comparisons test, p=0.0003, df = 264, n=23) and days 5, 6 and 8 post differentiation (two-way ANOVA with sidak’s multiple comparisons test, p>0.0001, df = 264, n=23).

A comparison of phenotype scores between *IGHMBP2*-KD and control iMN lines revealed that knockdown of a subset of genes exhibited reduced impact on survival in the *IGHMBP2*-KD background compared to control iMNs (50 genes, Fig. 5B). This suggests that *IGHMBP2 LOF* may drive toxicity in iMNs through biological processes implicated by these sgRNA targeted genes, as knockdown of genes within these pathways resulted in diminished toxicity with *IGHMBP2*-KD. The buffering of these sgRNA by *IGHMBP2*-KD was not detected in iCN, suggesting that this impact was neuronal subtype specific. GO analysis identified tRNA, rRNA, RNA splicing and ribonucleoprotein (RNP) complex biogenesis-related biological processes amongst the top five pathways buffered by the knockdown of IGHMBP2 in iMNs, implicating nuclear/nucleolar-dependent translational defects in iMN survival (Fig. 5C, Table S6). If *IGHMBP2*-LOF induces toxicity through disruption of RNP/tRNA/rRNA-related pathways, the knockdown of genes within pathways involved in tRNA/rRNA/RNP function would likely have diminished additional impact on viability in *IGHMBP2*-KD cells, supporting a model in which RNP/tRNA/rRNA dysregulation contributes to motor neuron degeneration in SMARD1 and CMT2S. To complement this finding, longitudinal imaging analysis was used to evaluate the impact of *IGHMBP2* LOF on iMN and iCN survival. A reduction in survival was detected with knockdown of *IGHMBP2* in iMN but not iCN at day 4 post differentiation (p=0.0003) and days 5, 6 and 8 post differentiation (p>0.0001) (Fig. 5D).

### Functional restoration of dysregulated pathways in treated intron 8 variant iMNs

Following successful restoration of IGHMBP2 protein for two of three intron 8 variants using the ASO-mediated splice correction, we next investigated whether the identified downstream cellular pathways were also restored in patient iMNs. Transcriptomic differential expression analysis was performed to assess pathway-level changes. Principal component analysis (PCA) revealed substantial separation between patient-derived and control samples, indicating marked transcriptomic differences (Fig S8A-D). To reduce false positives arising from genetic background effects, differential expression analyses were performed within patient-derived treatment groups rather than relative to control samples. Samples were stratified into “Successful” (c.1235+894C>A and c.1235+1076G>A) and “Unsuccessful” (c.1235+450G>A) categories, and comparisons were conducted between treatment conditions (Fig. S9). Few genes were differentially expressed across comparisons, suggesting no direct role for IGHMBP2 in gene expression (Fig. 6A; Fig. S10; Tables S7-12).

**Figure 6:**
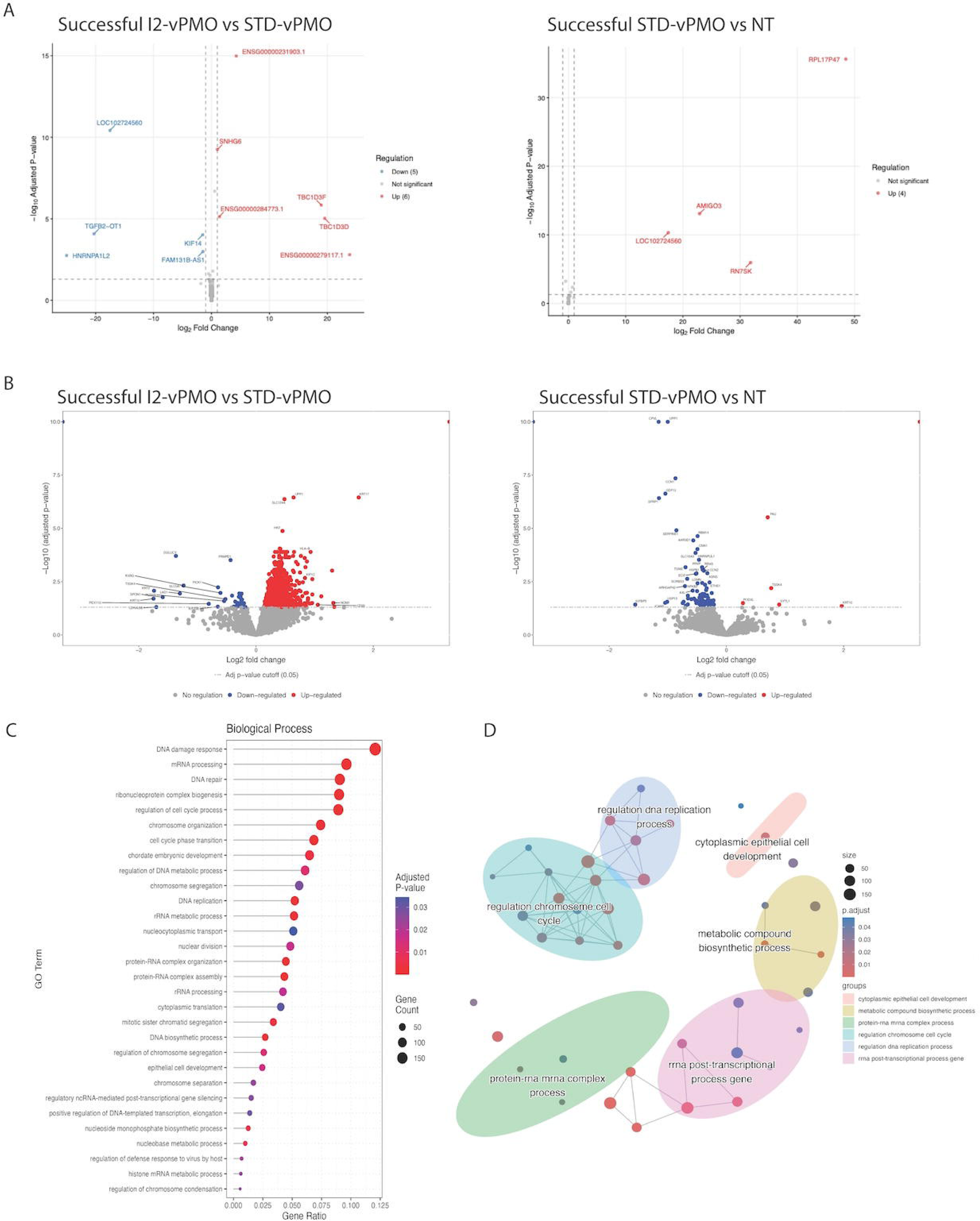
Successful treatment of patient derived iMNs with I2-vPMO reveals restoration of IGHMBP2 function. Comparison of treatment conditions within successfully treated samples (c.1235+894C>A and c.1235+1076G>A) versus unsuccessfully treated samples (c.1235+450G>A) A) Transcriptomic analysis reveals few differentially expressed genes between any treatment conditions B) proteomics analysis reveals strong protein upregulation in the successfully treated iMNs (I2-vPMO) compared with STD-vPMO C) Gene ontology analysis of these upregulated proteins highlights pathways of rRNA processing, RNA splicing and RNP biogenesis included in the top 30 categories. D) semantic analysis of top 50 categories organizes pathways into 6 primary groups including rRNA processing, RNP biogenesis, actin pathways and mRNA-protein interactions.

Given evidence from a CRISPRi screen linking IGHMBP2 to translational regulation, proteomic analysis was performed to assess whether *IGHMBP2* restoration resulted in changes at the protein level instead. The same comparison framework used for transcriptomic differential expression analysis was applied to identify differentially abundant proteins (Fig. S8 and S11A-G). Comparison of successfully treated I2-vPMO samples with STD-vPMO samples revealed widespread upregulation of proteins (n=1664), whereas only modest changes were observed in the other comparison groups (Fig. 6B, Fig. S12A-C, Tables S13-17).

Gene ontology analysis of differentially abundant proteins identified amongst the top 30 upregulated biological process pathways to include cytoplasmic translation, RNA splicing, rRNA processing, and ribonucleoprotein biogenesis (Fig. 6C, Table S18). Notably, pathways involved in DNA replication and DNA damage response were also enriched, consistent with previously reported functions of *IGHMBP2* (*11*). To consolidate related pathways, the top 50 gene ontology terms were assessed for semantic similarity and grouped into broader functional categories (Fig. 6D, Fig. S12D-E). The dominant themes included rRNA processing, ribonucleoprotein complexes, helicase activity, DNA/RNA binding and actin-related pathways (Fig. 6D, Fig. S12D-E).

Recent studies have shown that IGHMBP2 binds ribosomes in the cytoplasm and selectively regulates translation of proteins involved in mRNA homeostasis without globally altering translation in HeLa cells (*8, 11, 19, 20*). To assess whether a similar cytoplasmic based translational mechanism operates in iMNs, protein abundance was correlated with RNA abundance to identify proteins with concordant or discordant RNA–protein relationships (Tables S19-23). Enrichment of discordant proteins would be consistent with direct ribosomal translational regulation by IGHMBP2. A large upregulation of protein abundance is seen in successfully treated iMNs, but only a small subset is discordant with RNA (n=426, Fig 7A-B, Fig S13A-C). To characterize proteins potentially regulated at the translational level by IGHMBP2, statistically significant concordant and discordant RNA–protein pairs were annotated using gene ontology analysis (Fig. 7D-F, Table S24). Discordant proteins are predominantly enriched for pathways involved in RNA homeostasis and nucleocytoplasmic export, consistent with a previous study identifying nucleocytoplasmic export through THO complexes as a key pathway regulated by *IGHMBP2* (*19*) (Fig. 7D-F).

**Figure 7:**
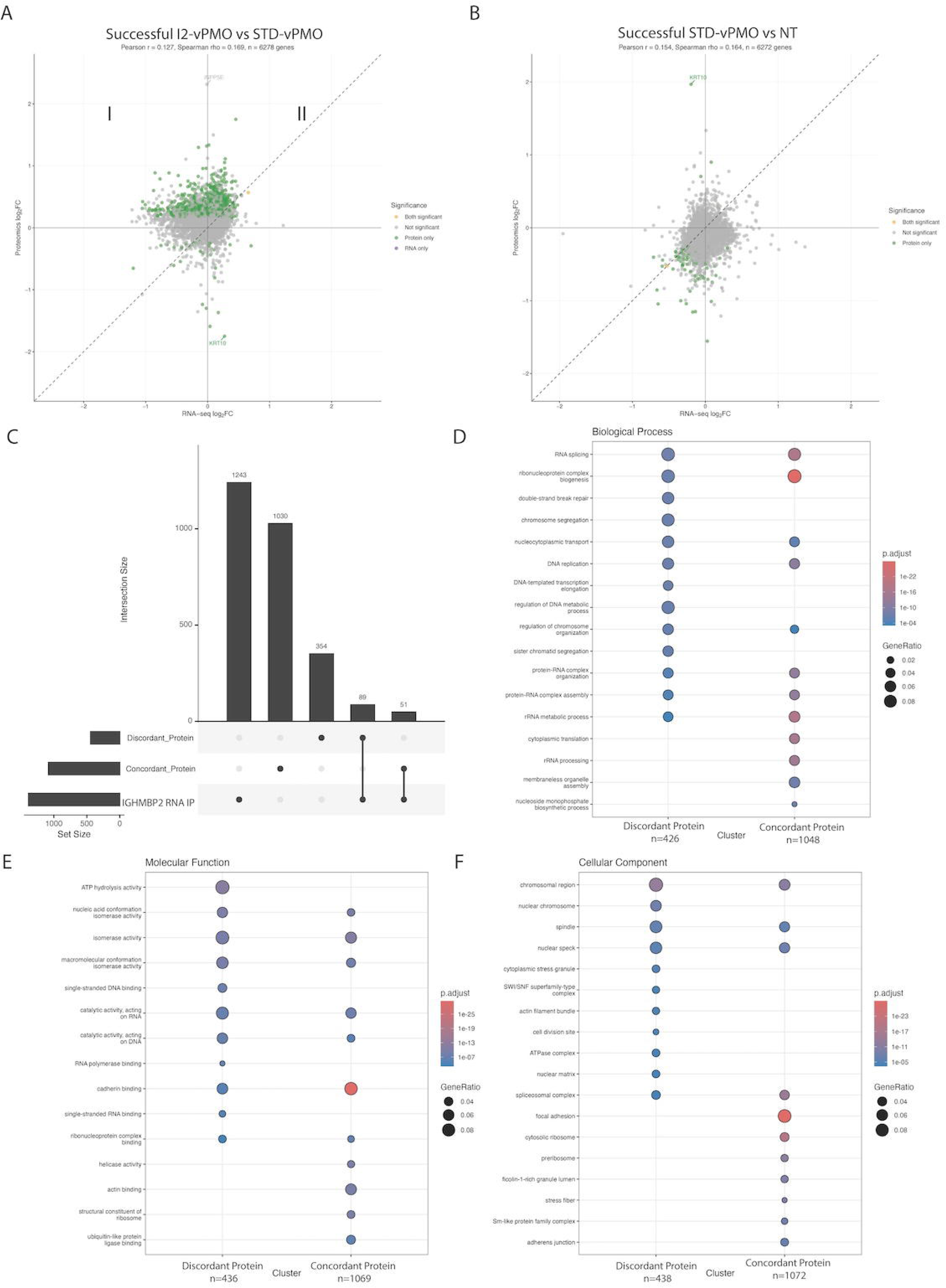
Protein-RNA abundance correlation highlights protein upregulation with successful I2-vPMO treatment discordant from RNA. A) Correlation of successful treated samples I2-vPMO to STD-vPMO reveals a leftward and upward shift which can be split into two quadrants. In quadrant I, protein abundance is upregulated discordant from RNA abundance, and in quadrant II, protein abundance is upregulated in concordance with upregulated RNA. B) no leftward and upward shift is seen when comparing successfully treated STD-vPMO to NT conditions C) enrichment analysis of discordant protein set with RNA IP reveals strong enrichment (89 overlapping genes; enrichment fold 28.11; Fisher’s exact test, p < 2 × 10⁻¹⁶) and analysis of concordant set reveals smaller enrichment (51 overlapping genes; enrichment fold 6.6; Fisher’s exact test, p < 2 × 10⁻¹⁶). D-F) Gene ontology significant categories in discordant protein set vs concordant protein set reveals pathways of mRNA homeostasis and nucleocytoplasmic export in the discordant (and concordant sets), but not pathways of rRNA processing or ribosome biogenesis, which appear only in the concordant set.

To assess whether IGHMBP2 binds to mRNA of upregulated proteins, RNA immunoprecipitation was performed using an antibody against *IGHMBP2*. The top 10% of enriched transcripts were defined as a high-confidence *IGHMBP2* RNA-binding set. Enrichment analysis was then used to compare overlap between this binding set and the discordant and concordant protein groups. The discordant protein set showed strong enrichment for IGHMBP2-bound transcripts (89 overlapping genes; enrichment fold 28.11; p < 2 × 10⁻¹⁶), whereas the concordant set showed substantially lower enrichment (51 overlapping genes; enrichment fold 6.6; p < 2 × 10⁻¹⁶; Fig. 7C, Table S25). Notably, *THOC2* was included among the discordant proteins overlapping with IGHMBP2-bound RNAs, consistent with prior reports that *IGHMBP2* binds *THOC2* and enhances its translational efficiency (Table S25) (*19*).

### Decreased local translation is a cellular phenotype of SMARD1 iMNs

Whether translation is globally or locally affected in *IGHMBP2* LOF remains uncertain (*18, 32, 33*). Local translation is critical in motor neurons, and prior studies have reported reduced β-Actin local translation in growth cones of *IGHMBP2*-deficient mouse primary motor neurons (*32, 33*). Analysis of growth cones in iMNs from the c.1235+1076G>A patient (P8) and *IGHMBP2* KO lines at 7 days post-differentiation showed significantly reduced β-Actin intensity compared with controls (p < 0.0001; Fig. S14A-B). However, analysis of the cell bodies, reflecting global translation, showed no significant difference in β-Actin intensity (p=0.1198; Fig. S14C). This suggests that translational defects due to IGHMBP2 LOF may lead to an axonal growth specific vulnerability. However, the precise relationship between IGHMBP2 and local translation remains unclear and warrants further investigation.

## Discussion

In this work we report twelve unrelated patients of varied ethnic backgrounds with rare deep intronic variants in intron 8 of *IGHMBP2* (c.1235+450G>A, n=5; c.1235+894C>A, n=1; and c.1235+1076G>A, n=6) in compound heterozygosity with recognized likely pathogenic variants in *IGHMBP2,* presenting with phenotypes consistent with SMARD1 or CMT2S. Short- and long-read RNA sequencing performed in patient-derived induced motor neurons (iMNs) demonstrated that long-read sequencing from a disease-relevant cell type was required to fully resolve pathogenic splicing. This sequencing revealed that all three variants generate overlapping pseudoexons through distinct novel acceptor sites that converge on a shared cryptic donor and are subsequently degraded. Clinically, patients exhibited comparatively milder phenotypes than typically reported for individuals with biallelic *IGHMBP2* variants, including delayed respiratory involvement in two cases (*5–8*). Consistent with early observations that disease severity correlates with the amount of preserved full-length and functional *IGHMBP2* protein, variants in intron 8 were associated with residual canonical splicing (approximately 30–75%), likely permitting limited production of functional protein and supporting a phenotypic continuum from SMARD1 to CMT2S (*6, 8, 34, 35*). Accordingly, patients harboring deep intronic *IGHMBP2* variants may warrant longitudinal respiratory surveillance despite initially milder presentations.

Splice prediction tools, including SpliceAI and Pangolin, failed to accurately predict the splicing consequences of deep intronic variants in *IGHMBP2* intron 8, underscoring a broader challenge in motor neuropathy diagnostics, where molecular diagnostic rates remain low (∼30%) despite extensive gene discovery (*36*). Identifying intronic regions predisposed to aberrant splice activation may therefore improve diagnostic yield (*37–41*). *IGHMBP2* intron 8 exhibits low-level cryptic donor and acceptor usage in control iMNs, and Ensembl annotations include non–protein-coding transcripts incorporating these pseudoexons. Whether this region contains a poison exon or is merely a yet unrecognized alternative coding exon remains unclear (*37, 42*). Rare variant-created pathogenic pseudoexons are often located in regions where low-level cryptic splicing is observed in control cells, suggesting that genomic features of the region (such as poison-exon features) confer susceptibility to pathogenic splice activation (*37, 38, 43*). Long-read RNA sequencing in disease-relevant cell types, such as motor neurons, may enable systematic identification of these predisposed regions and clarify how local genomic features, including repetitive elements, contribute to splice vulnerability, evasion of standard detection methods, and inform the interpretation of deep intronic variants (*44, 45*).

Antisense oligonucleotide (ASO) development for rare disease is resource-intensive, and variant-specific designs limit scalability. A previously reported ASO targeting the *IGHMBP2* c.1235+894C>A variant was directed against a variant-specific acceptor site and is therefore not generalizable to variants that are different (*10*). In contrast, the variant class-directed ASO strategy evaluated here corrected aberrant splicing and restored IGHMBP2 protein for the c.1235+894C>A and c.1235+1076G>A variants, but not c.1235+450G>A, likely reflecting limitations of a single-ASO approach across pseudoexons of differing lengths. Nonetheless, the recurrence of distinct intronic variants on shared cryptic splice sites—observed in *IGHMBP2* and at least twelve additional loci—suggests that such unified splice-corrective strategies may have broader therapeutic applicability for similar clusters of variants (*38, 43*).

Although this work demonstrates that increasing *IGHMBP2* bioavailability can attenuate disease phenotypes, the basis for clinical heterogeneity between severe SMARD1 and milder CMT2S -- particularly in patients with biallelic missense variants -- remains enigmatic. *IGHMBP2* consists of four primary domains, the largest which comprises a helicase ATP-binding domain. The C-terminus includes a Zinc Finger and R3H domain. Most pathogenic variants localize to the helicase ATP-binding domain, implicating disrupted pre-rRNA and tRNA processing, whereas comparatively few affect the C-terminus, which mediates ribosomal binding (*1, 3, 5, 8, 46–48*). These observations support a model in which IGHMBP2 participates in multiple molecular pathways, with differential disruption contributing to phenotype severity (*11, 47, 48*). Elucidating IGHMBP2 interaction partners will therefore be essential to define genotype–phenotype relationships, identify pathway-specific vulnerabilities and provide candidates for modifiers of IGHMBP2 phenotypic severity.

Dysregulation of translation is increasingly recognized as a key driver of axonal neurodegeneration (*49–51*). Our CRISPRi screen identifies tRNA, rRNA, and RNP processing pathways as particularly vulnerable in motor neurons, supporting a critical nuclear/nucleolar role for IGHMBP2 in maintaining translation through pre-rRNA and tRNA processing. This is consistent with findings in the *nmd* mouse model, where tRNA^Tyr^ supplementation and modulation of activator of basal transcription 1 (*ABT1*)—a regulator of pre-rRNA processing—alter disease severity, and where mutations in *IGHMBP2* impair *ABT1* binding and correlate with reduced enzymatic function (*11, 22, 24, 48, 52–54*). In contrast, IGHMBP2 also functions in the cytoplasm, where it associates with ribosomes via its C-terminus and regulates translation (*17–20*). In K562 cells, *IGHMBP2* knockdown causes mild global translational downregulation (*18*), while in HeLa cells it selectively impairs translational efficiency of proteins with structured, GC-rich 5′ UTRs, including those involved in mRNA homeostasis (*19*). In primary motor neurons from *nmd* mice, translational defects are spatially restricted to β-actin in growth cones rather than cell bodies (*32*). Consistent with these findings, we observed reduced β-actin intensity at growth cones but not in the cell bodies in *IGHMBP2* KO and c.1235+1076G>A patient-derived iMNs. Given that ribosomes are present along axons and that motif-dependent ribosomal proteins are locally translated in growth cones to support axonal translation (*51, 55, 56*), IGHMBP2 may have a role in regulating localized, motif-dependent translation, thereby impacting axon growth. Together, these findings support a model in which IGHMBP2 maintains translational homeostasis through dual functions in nuclear/nucleolar RNA processing and cytoplasmic, localized translation, with disruption of either axis contributing to motor neuron vulnerability.

Evidence for IGHMBP2’s dual nuclear and cytoplasmic roles is observed in successfully treated intron 8 patient-derived iMNs, where upregulated proteins segregate into concordant and discordant sets. The discordant protein set is enriched for IGHMBP2 binding targets and gene ontology analysis revealed pathways related to mRNA homeostasis. In contrast, the concordant set uniquely included ribosome biogenesis, rRNA processing and cytosolic translational pathways. Together, these findings suggest that IGHMBP2 may enhance translational efficiency both directly—through ribosome association—and indirectly, by restoring rRNA processing and improving the function of RNP-dependent complexes, ultimately permitting downstream increases in ribosome abundance. Notably, while these proteomic results are consistent with prior reports implicating IGHMBP2 in nucleocytoplasmic export and mRNA homeostasis, rRNA processing and RNP pathways have not been detected in previous profiling studies (*18, 19*). This discrepancy highlights the importance of disease-relevant cellular models, which can uncover mechanistic insights that are obscured in non-relevant systems.

Importantly, the treated patient-derived iMN transcriptomic and proteomic datasets developed here enables in-vivo biomarker discovery, which will be critical as *IGHMBP2* gene replacement and antisense oligonucleotide therapies advance toward clinical application (*10, 13, 15, 16*). With slowly progressive diseases such as CMT2S, clinical outcome measures may not detect meaningful change at earlier timepoints following treatment (*57, 58*). Imaging or serum biomarkers represent an attractive alternative to evaluate target engagement and establish therapeutic efficacy (*57, 58*). Further work with a larger cohort of *IGHMBP2* patients and more diverse phenotypes is warranted to explore and validate potential biomarkers.

A limitation of this study is the inability to detect rescue of tRNA metabolism or integrated stress response (ISR) pathways following *IGHMBP2* correction, despite prior work implicating aminoacyl-tRNA synthetase dysfunction and ISR activation in inherited neuropathies and *IGHMBP2* deficiency (*18, 49, 50, 59–78*). Although CRISPRi screening identified *IGHMBP2* as a mediator of tRNA metabolism, transcriptomic and proteomic analyses of treated intron 8 iMNs did not reveal normalization of these pathways, and ISR activation was not observed in the proteomics dataset. This may reflect technical limitations of mass spectrometry, including limited sensitivity for low-abundance or transient stress signaling, or the absence of targeted ISR interrogation. Alternatively, *IGHMBP2* restoration may preferentially rescue downstream translational capacity without fully normalizing upstream tRNA charging or stress signaling in our treatment timeframe. Targeted analyses will be required to clarify the relationship between *IGHMBP2*, tRNA metabolism, and ISR activation in motor neuron vulnerability.

## Conclusion

In conclusion, long-read transcriptomic analysis helped uncover a deep intronic pathogenic hotspot in *IGHMBP2*, enabling splice-corrective intervention and reinforcing the value of long-read sequencing in disease-relevant cell types to inform diagnosis, therapeutic strategy, and biomarker discovery in motor neuropathies.

## Methods

### Patient Recruitment

Patients were referred by their neurologist or geneticist. Written informed consent and age appropriate assent for study procedures were obtained by a qualified investigator [protocol 12-N-0095 approved by the National Institute of Neurological Disorders and Stroke, National Institutes of Health Institutional Review Board (IRB); Siriraj IRB (COA no. Si 501/2017 and 601/2023); University of Pennsylvania IRB under protocol number 832955 (with support through NIH U54NS065712); National Human Genome Research IRB under research protocol 15-HG-0130; Nationwide Children’s Hospital under study IRB11-00215]. Medical history, clinical evaluations and muscle biopsy were obtained as part of the standard diagnostic examination. Samples for research-based testing, including skin, blood and muscle biopsy samples, were obtained using standard procedures.

### Neuron differentiation and ASO treatment

Insertion of transcription factors NGN2, LHX, and ISL1 (NIL) was completed to promote lower motor neuron differentiation (*26*). iPSCs were co-transfected using the PiggyBac system with N50-PB-tri-TO-NIL-BFP-puro plasmid (addGene #172113) and K13-EF1a-transposase as previously described (*79*). BFP positive cells were selected with 10ug/ml puromycin. NIL+ cells were expanded and cryopreserved at passage +2 post selection. Cells were thawed and used for experiments within 2 passages to maintain high differentiation efficiency. 1x10^6^ NIL containing iPSCs were differentiated on a 10cm dish coated in Matrigel (Day 0) as previously described (*80*), with the addition of 1 µM Uridine and Fluorodeoxyuridine (FDU) on day 2. Three 10 cm dishes were combined and replated on Day 3 to one Matrigel coated 6 well plate with neuronal maintenance media (NMM) (50% DMEM/F12, 50% Neurobasal Media with 1x GlutaMAX (Gibco, 35050061), 1x B-27 Supplement (Gibco, 17504044), 1x N-2 Supplement (Gibco, 17502048) and 1x MEM Non-Essential Amino Acids (Gibco, 11140050)) supplemented with 2ul/ml doxycycline, 10µM ROCK-inhibitor, 10ng/ml BDNF (Gibco, PHC7074), 10ng/µl GDNF (Gibco, 450-10-10UG), 10ng/µl CNTF (Gibco, 450-13-20UG), 10ng/ml NT-3 (Gibco, 450-03-10UG), 1µM FDU, 1ug/ml laminin (Gibco, 23017015). For ASO treatments, a one-time 1µM dose of either the custom IGHMBP2 Vivo-Morpholino or a Vivo-Morpholino standard control (Gene Tools, LLC) was supplemented to the culture medium, four hours after cell replating on day 3. Half of the NMM was removed and replaced on Day 6. Cells were collected on day 7.

### Western blot

30ug of protein was normalized to 40ul with RIPA buffer with 4xSB dye. Samples were heated at 95°C for 5 minutes, loaded into Novex 10% tris-glycine gel (Invitrogen, XP00105BOX) with Novex 1x tris-glycine SDS running buffer (Invitrogen, LC2675) and separated for 90 minutes at 125 volts. Protein was transferred to a PVDF 0.45 pore membrane using Novex 1x tris-glycine transfer buffer (Invitrogen, LC3675) with 10% methanol at 25V for 90 minutes. Membranes were stained with Ponceau to confirm transfer, then blocked with 5% milk in 1% TBS-tween (TBS-T) for 1 hour. Primary antibodies were incubated at 4°C overnight (IGHMBP2 anti mouse 1:1000 (Millipore clone 11-24, MABE 162), rabbit anti GAPDH 1:2000 (Cell Signaling Technologies, 5174)) in 5% milk at 4°C. Samples were rinsed 3x in TBS-T and then incubated with secondary 1:10000 (Peroxidase-conjugated AffinPure goat anti mouse and goat anti rabbit IgG from Jackson ImmunoResearch 115-035-044 and 111-035-003) for 1 hour at room temperature. Membranes were rinsed 3x in TBS-T, incubated for 1 min in ECL-Select western blotting detection reagent (Cytiva, RPN2235) and imaged using the chemidoc with 2-minute exposure.

### Long read RNAseq alignment

RNA extracted from cells were prepared with either size selection or probe pull down to increase reads over *IGHMBP2* and sequenced on PacBio Revio (supplemental Methods). HiFi reads were demultiplexed and adapters removed with Lima (v2.0.3). Full-length, non-concatemer (FLNC) reads were generated from raw subreads, and transcript isoforms were clustered using the Iso-Seq3 pipeline (v4.3.0). The resulting high-quality polished consensus transcripts were aligned to the GRCH38 reference genome (Gencode, version 45) using pbmm2 (v1.16.0) with the --preset ISOSEQ option. Transcript redundancy was removed using isoseq3 collapse to generate a final GFF/GTF file. Pigeon (v1.4.0) was used to classify and annotate transcripts.

### Long read phasing

Raw read files were subset with seqtk (v1.5-r155) to reads spanning exon 8 - 9 of *IGHMBP2*. Reads were haplotyped and BAM files subsequently tagged using IsoPHASE (v.29.0.0) tools (https://github.com/Magdoll/cDNA_Cupcake/wiki/IsoPhase:-Haplotyping-using-Iso-Seq-data#need). BAM files were then split by haplotype tag using Picard FilterSamReads (v4.0.10) and indexed. Sashimi plots were generated with ggSashimi (*81*). Allele specific Percent Spliced In (asPSI) values were manually calculated for each sample by dividing the splice site of interest by the total splice junctions from the shared starting point. Control samples were not phased due to insufficient haplotypes present in the samples.

### Short read RNAseq and differential expression analysis

Samples were enriched for mRNA containing Poly A tails using the NEBNext polyA mRNA magnetic isolation module (New England Biolabs, Ipswitch,MA). From this, strand-specific, dual unique indexed libraries were made using the NEBNext® UltraExpress RNA Library Prep Kit for Illumina® (New England Biolabs,Ipswich, MA). Libraries were sequenced on an Illumina NovaSeq 6000 using 150bp PE reads.

Quality control of FASTQ files were assessed with FastQC (v0.12.1). BAM files were generated using the GTEXv10 pipeline (https://github.com/broadinstitute/gtex-pipeline) using reference genome GRCh38 (Gencode, version 45). Briefly, FASTQ files were aligned using STAR (v2.7.10a) with custom splice junctions specified (--sjdbFileChrStartEnd) and optical duplicates marked by Picard Mark Duplicates (v3.3.0) (*82*). Alignment metrics were assessed with RNA-SeQC (v2.4.2) (*83*). Sashimi plots were generated using ggSashimi (*81*). Percent Spliced In (PSI) values were quantified using MAJIQ (v2.5) with build command customized for --min-denovo 2 and comparisons between conditions using deltapsi (*84*). Gene counts were produced with Salmon (v1.10.1) (*85*). Count matrices were imported into RStudio (R v4.5) with tximport. PCA analysis was performed using both DESseq2 and prcomp function from stats package (v.4.5.2) and graphed with plotly (v.4.11.0). Differential expression analysis was performed with DESseq2 after collapsing replicates (n=3 per condition), using alpha 0.01 and false discovery rate (FDR) 0.05 (*86*). Ensembl IDs were converted to gene symbols using AnnotationDbi and org.Hs.eg.db packages. Volcano plots were generated by shrinking log2fc with type “ashr” and plotted using ggplot2 (*87*).

### Mass spec and differential proteomics analysis

Samples were prepared for mass spectrometry (Supplemental Methods). Data were acquired in data-independent mode (DIA) on an Orbitrap Ascend mass spectrometer (Thermo Fisher Scientific) coupled with a Vanquish Neo HPLC (Thermo Fisher Scientific). Spectronaut 20.3 software was used to analyze DIA data with the directDIA analysis method. Data were searched against Sprot Human database. Trypsin/P was set as a specific enzyme with 2 max missed cleavages. Carbamidomethyl on cysteine was a fixed modification. Acetyl (Protein N-term and Oxidation on methionine) were set as variable modifications. FDRs for PSM, peptide and protein level were 1%. MS2 abundances were used for protein quantitation. Cross-run normalization was performed. Protein abundances were exported to MSstats for analysis.

Spectronaut output was imported into RStudio and prepared with MSstatsConvert (v.1.20.0) function SpectronauttoMSstatsFormat. Protein abundance was quantified and normalized with MSstats (v.4.18.0) with flags –censoredINT =0, featureSubset = “highQuality”, remove_uninformative_feature_outlier = TRUE, and min_feature_count = 2 (*88*). PCA analysis was performed using protein abundances from MSstats with prcomp and graphed using plotly. Differential protein analysis was performed using the groupComparisons function of MSstats with FDR 0.05. Protein names were converted to gene symbols using AnnotationDbi and org.Hs.eg.db packages. Volcano plots were generated using the groupComparisonPlots function in MSstats.

### RNA-Protein correlation and concordance analysis

Log2 fold changes from both RNAseq and Proteomics analysis were subset to contain proteins/genes captured by both methods. Correlation was performed using in house scripts. Concordance was estimated using the sign function on log2 fold change values from both RNA and protein, with concordance ascribed when the output was equal and discordance ascribed when the output differed between RNA and protein.

### Enrichment Analysis

Enrichment analysis of the top ten percent of enriched IGHMBP2 binding sites was performed for both concordant and discordant protein datasets (see Supplemental Methods for RNA IP methods). Gene set enrichment was assessed using a one-sided Fisher’s exact test to determine whether genes in set A were overrepresented in set B. Gene lists were deduplicated, and overlap was defined as their intersection. A 2×2 contingency table was constructed based on the number of genes in each set and a background gene set of all human genes. Expected overlap was calculated as (*η*_*A*_ × *η*_*B*_)/*N*, and fold enrichment as the ratio of observed to expected overlap. Odds ratios and p-values were reported, with significance defined as p < 0.05.

### Gene ontology analysis

Over representation analysis of differentially expressed genes and proteins was performed using ClusterProfiler (*89*). Briefly, lists of differentially expressed genes/proteins in gene symbols were compared to background genes/proteins (all genes/proteins quantified) using the hypergeometric test with functions enrichGO or clusterCompare (fun= “enrichGo”) with pValue and qValue set to 0.001, padjust method set to Benjamini-hochberg. Simplification of terms was performed with simplify function in cluster profiler, cutoff=0.7. Lolliplots and dotplots were made with ggplot2. Emmaplots were made by first performing semantic similarity analysis using pairwise_termsism function, then graphed using emmaplot function with node_label = “group”.

### Survival CRISPRi screen

iPSCs containing the dCas9 and NGN2 or hNIL cassettes were expanded to 50 million cells and transduced with a whole genome guide library (hCRISPRi-v2) to a titer of approximately 80% in E8 Medium containing ROCK inhibitor (Supplemental Methods) (*90*). The following day the media was changed to E8 without ROCK-I. Two days after transduction the cells were split and iPSC were treated with 11ug/mL puromycin in E8 medium with ROCK-I to select for iPSC expressing sgRNA. The following day the medium was changed and after two days the cells were passaged and transduced with control or IGHMBP2 sgRNA lentiviral samples containing blasticidin resistance. Two days after transduction, the cells were treated with 15ug/mL blasticidin. The following day the media was changed to E8 media only and then two days after, 200 million cells from either the IGHMBP2 or control sgRNA NIL and NGN2 treated samples were collected at baseline (Day 0). The remaining cells were plated on PEI coated dishes at a density of 10 million cells per 15cm dish. Between 150-200 million cells were collected at each day of collection (Days 3 or 14) for each cell type (NIL or NGN2) and with each sgRNA (Control or IGHMBP2). The differentiation was started at day 0 in Neuronal Induction Media (NIM) (supplemental methods) with doxycycline (2ug/mL) and ROCK-I (10uM). On day 2, half of the NIM media was removed and replaced with Neuronal Maintenance Media (NMM) including supplements indicated above. One-half of the media was replaced every 3 days. Two biological replicates were performed for each time point, cell line, and sgRNA yielding a total of 24 different genomic DNA libraries for sequencing. Genomic DNA was extracted (see supplemental methods) and the sgRNA containing amplicons were PCR amplified in 10ug genomic DNA reactions containing 1uM with one of the following index primers for each of the 24 libraries: (see Table S26) and 1uM of the following reverse primer

CAAGCAGAAGACGGCATACGAGATCGACTCGGTGCCACTTTTTC. Sequencing was performed with the custom primer

GTGTGTTTTGAGACTATAAGTATCCCTTGGAGAACCACCTTGTTGG, and the illumina index sequencing primer GATCGGAAGAGCACACGTCTGAACTCCAGTCAC on a NovaSeq S2 flow cell with 8% PhiX spike in. Data analysis was performed using the MAGeCKFlute pipeline to assess biological functions and pathways of genes enriched or depleted in the cultured neurons and statistical analysis was performed by analyzing the phenotype score (day of culture – baseline) and FDR.

### Survival Imaging

Ten thousand iPSCs expressing >95% control or *IGHMBP2* sgRNA were plated in each well in a 384-well plate in 100uL Induction Media with doxycyclin (2ug/mL) and ROCK-inhibitor (10uM). Wells along each edge of the plate were filled with 150uL PBS to maintain humidity. A total of 22 wells were imaged for each cell line.

Images were acquired on days 2, 3, 4, 5, 6, and 8 post-induction on a Nikon Ts2 inverted microscope using an automated stage at 37°C and 5%CO_2_ using Nikon Elements software. On days 2 and 6, 50% of media was removed and replaced with NMM media supplemented with 10ng/mL NT3, 10ng/mL BDNF, 1ug/mL Laminin, and 2ug/mL doxycycline. Image analysis was performed with Cell Profiler to quantify the number of cells expressing *IGHMBP2* sgRNA. Survival was analyzed as fraction from baseline cell number.

## Data Availability

Datasets generated in this manuscript are available on dbGAP phs001322.v4.p1 or on reasonable request from authors.

## List of Supplementary Materials

Supplemental Figures

Supplemental Tables

Supplemental Methods

Supplemental Case Reports

## Acknowledgments

### Declarations

This research was supported by the Intramural Research Program of the National Institutes of Health (NIH) [grant 1ZIANS003129 and ZIANS009455]. The contributions of the NIH author(s) are considered Works of the United States Government. The findings and conclusions presented in this paper are those of the author(s) and do not necessarily reflect the views of the NIH or the U.S. Department of Health and Human Services. OS was supported by Health Systems Research Institute (HSRI 67-104). FV, EC, AC, KT, KP, KL were supported by the Indiana University Grand Challenge Precision Health Initiative. DH was supported in part by NIH 2U54NS065712. TC and MK were supported by NIH Common Fund, NIH/NHGRI grant UO1HG007674. APM, DCK, AMC, ARM, and RS received funding from the Nationwide Foundation Innovation Fund for the patient studies.

### Data and Materials Availability

Datasets generated in this manuscript are available on dbGAP phs001322.v4.p1.

### Author contribution

SS: Conceptualization, Investigation, Formal analysis, Writing – original draft.

AN: Formal analysis.

RO: Clinical data curation, Project administration.

SD, TC, MK, JB, DH, FV, EC, AC, KT, KP, KL, Alayne M, DK, AMC, RS, Anthony M, PK, OS, KS, DBG, WM, AS, JP, AYM, ARH, KRC, Sarah N: Clinical Evaluation, Clinical Data curation.

VB, AW, EM, Shreya N, GH, BJ, KJ, JJB, JM, IS, AP, KP, YL, SK-H: Investigation, Formal analysis.

DA, MW: Conceptualization, Methodology.

CG, CB: Conceptualization, Methodology, Funding acquisition, Supervision.

All authors: Writing – review & editing.

## Acknowledgments

We thank Jizhong Zou from the NHLBI iPSC core for helping to generate the IGHMBP2-KO iPSC line. We thank the NHLBI and NINDS iPSC cores for producing control and patient derived iPSC lines used in this study. We thank Ben Weisburd of Broad Institute for insights into repetitive elements surrounding intron 8. This work utilized the computational resources of the NIH HPC Biowulf cluster (https://hpc.nih.gov). With gratitude to all patients and families for participating in the study. Editing of this manuscript was supported with AI technology (NIH CHIRP ChatGPT v5.4).

## Supplementary Materials

Figure S1: Validation of iPSC derived iMNs. A) karyotyping of iPSCs for c.1235+450G>A, c.1235+894C>A and c.1235+1076G>A lines revealed no genomic rearrangements. B) iMNs stain positive for motor neuron markers HB9 (red), β-tubulin (green) at five days post differentiation.

Figure S2: Presence of biallelic variants in iGHMBP2 allow for phasing of long reads by haplotype. A) long reads for both maternal and paternal alleles for c.1235+450G>A, B) for c.1235+894C>A and C) for c.1235+1076G>A lines.

Figure S3: Initial testing of I2-PMO on c.1235+1076G>A fibroblast lines show dose response reduction in *IGHMBP2* pseudoexon bands. A) RT-PCR demonstrating reductions of pseudoexon containing band with 200nM-20uM dosing of I2-PMO treatment. STD-PMO (standard control) does not show reduction in pseudoexon band. B) normalization of pseudoexon or wildtype *IGHMBP2* bands to GAPDH and to untreated conditions demonstrates statistically significant increase in wildtype band with I2-PMO treatment at concentrations of 200nM and greater (two-way ANOVA with Tukey’s multiple comparisons test, df = 34, p < 0.0001, n=3 replicates per condition).

Figure S4: I2-vPMO treatment in patient derived iMNs does not raise total *IGHMBP2* mRNA levels. Normalized *IGHMBP2* counts do not increase significantly with I2-vPMO treatment compared to STD-vPMO or NT conditions for any cell line.

Figure S5: Long read RNA-seq of successfully treated variants complement short-read RNA-seq. A) I2-vPMO treatment in c.1235+1076G>A iMNs increases canonical splicing of exon 8 to exon 9, and no intronic usage is observed. B) A SNP present in exon 13 (black arrow, green SNP) phases with the c.1235+1076G>A allele. In I2-vPMO treated cells, the two reads with canonical splicing come from the intron 8 variant allele. In the STD-vPMO and NT conditions, reads phasing with the c.1235+1076G>A variant are not present or present at low levels, suggesting efficient degradation of pseudoexon containing isoforms. The reads with intronic events captured in these conditions are from the other allele with a frameshift in exon 9. C) I2-vPMO treatment in c.1235+894C>A iMNs increases canonical splicing and completely prevents the 182bp pseudoexon splice event, which is observed in STD-vPMO and NT conditions.

Figure S6: CRISPRi screen methodology and validation. A) NGN2 and hNIL containing iPSCs show increase in neuronal specific markers after 48 hours of differentiation. B) RNA-seq after 48 hours of differentiation shows distinct gene expression profiles in NGN2 versus NIL containing iPSCs. C) schematic for sCas9-KRAB sgRNA mediated knockdown and CRISPRi gene silencing D) hiPSC-derived MN and CN CRISPRi experiment approach. Naive NGN2 and NIL cells containing dCAS9-KRAB are transduced with sgRNA library, differentiated and collected at Day 0, Day 3 and Day 14 post differentiation for survival analysis. Next generation sequencing completed to obtain sgRNA frequencies and MAGeCK-iNC pipeline used to generate phenotype scores. E) representative western blot confirmation of IGHMBP2 knockdown using sgRNA in iPSCs and iMNs. Data from two nontargeting sgRNA’s shown as control.

Figure S7: Comparison of motor neurons to cranial neurons. Comparison of phenotype scores between MNs (x-axis) and CNs (y-axis) for day 3 vs. day 0, and day 14 vs. day 0.

Figure S8: Principal component analysis (PCA) of intron 8 patient iMN RNA-seq data. 2D PCA scatterplot showing A) samples colored by treatment condition. B) samples colored by cell line status, patient (intron 8 variant), control or IGHMBP2-KO. Samples cluster by *IGHMBP2* mutation status rather than treatment. 3D PCA plots C) colored by treatment condition and D) colored by individual cell line, showing how samples cluster by cell line rather than treatment.

Figure S9: Schematic demonstrating comparison strategy used for omics analysis. Patient samples were compared to each other, split by response to I2-vPMO as determined by protein analysis. Controls were also compared to one another.

Figure S10: Transcriptomic analysis reveals few differentially expressed genes between conditions. A) comparison of unsuccessful patient iMNs I2-vPMO to STD-vPMO. B) comparison of unsuccessful patient iMNs STD-vPMO to NT. C) comparison of control iMNs I2-vPMO to STD-vPMO.

Figure S11: Principal component analysis (PCA) of intron 8 patient iMN proteomics data. A) 2D scatterplots colored by cell line and treatment condition, B) 3D plot colored by cell line and treatment condition, C) 2D scatterplots colored by treatment condition and D) in 3D. Clustering is observed when colored by patient status only E) 2D scatterplot and F) 3D plot. G) scree plot demonstrating most of the variance is explained by the first three principal components.

Figure S12: Proteomics analysis reveals upregulation of protein for successfully treated samples but not for other samples and conditions. A) few differentially abundant proteins for unsuccessful patient samples I2-vPMO vs STD-vPMO and B) for STD-vPMO vs NT. C) control samples show few differentially abundant proteins with I2-vPMO treatment vs STD-vPMO. D) semantic analysis of top 50 gene ontology categories for successfully treated I2-vPMO vs STD-vPMO samples, annotated by Molecular Function and E) Cellular Component.

Figure S13: RNA-Protein abundance correlation shows no leftward and upward shift for A) Unsuccessful samples I2-vPMO vs STD-vPMO, B) Unsuccessful samples STD-vPMO vs NT and C) Control I2-vPMO vs STD-vPMO.

Figure S14: Decreased local translation observed in c.1235+1076G>A iMNs. A) representative images of growth cones from c.1235+1076G>A, control (I2^+/+^) and isogenic IGHMBP2 KO (I2^-/-^) cell lines. F-actin staining was used to identify growth cones and cell bodies (not shown) in iMNs and β-Actin intensity was quantified within. B) quantification of β-Actin intensity across growth cones from each condition, growth cones were quantified from across three separate experiments (I2^+/+^ (n=59), I2^-/-^ (n=57), c.1235+1076G>A (n=44); Shapiro-Wilk test p<0.0001; Kruskal-Wallis test with Dunn’s multiple comparisons, p<0.0001). C) quantification of β-Actin intensity across cell bodies from each condition, cell bodies were quantified from across three separate experiments (I2+/+ (n = 37), I2−/− (n = 23), c.1235+1076G>A (n = 9); Brown–Forsythe test p = 0.0370; Welch’s ANOVA p = 0.1198). DPI = Days Post Induction

Table S1: Clinical Details of all 12 patients with intron 8 variants

Table S2: PSI calculations

Table S3: Probes used for *IGHMBP2* pull down long read RNAseq

Table S4: sgRNA sequences targeting *IGHMBP2* and non-targeting control

Table S5: CRISPRi screen phenotype scores across all comparisons performed

Table S6: CRISPRi screen phenotype scores in iMNs with dual treatment of either control or *IGHMBP2* sgRNA.

Table S7: Differential expressed genes for successful intron 8 iMN I2-vPMO vs STD-vPMO

Table S8: Differential expressed genes for successful intron 8 iMN STD-vPMO vs NT

Table S9: Differential expressed genes for unsuccessful intron 8 iMN I2-vPMO vs STD-vPMO

Table S10: Differential expressed genes for unsuccessful intron 8 iMN STD-vPMO vs NT

Table S11: Differential expressed genes for control iMN I2-vPMO vs STD-vPMO

Table S12: Differential expressed genes for control iMN STD-vPMO vs NT

Table S13: Differential expressed proteins for successful intron 8 iMN I2-vPMO vs STD-vPMO

Table S14: Differential expressed proteins for successful intron 8 iMN STD-vPMO vs NT

Table S15: Differential expressed proteins for unsuccessful intron 8 iMN I2-vPMO vs STD-vPMO

Table S16: Differential expressed proteins for unsuccessful intron 8 iMN STD-vPMO vs NT

Table S17: Differential expressed proteins for control iMN I2-vPMO vs STD-vPMO

Table S18: statistically significant pathways from gene ontology analysis for successful intron 8 iMNs I2-vPMO vs STD-vPMO

Table S19: RNA-protein correlation for successful I2-vPMO vs STD-vPMO

Table S20: RNA-protein correlation for successful STD-vPMO vs NT

Table S21: RNA-protein correlation for unsuccessful I2-vPMO vs STD-vPMO

Table S22: RNA-protein correlation for unsuccessful STD-vPMO vs NT

Table S23: RNA-protein correlation for control I2-vPMO vs STD-vPMO

Table S24: RNA-protein concordance estimates for successful I2-vPMO vs STD-vPMO

Table S25: RNA IP genes that intersect with discordant and concordant protein sets

Table S26: list of 24 primer sequences for CRISPRi genome sequencing

## Notes

### Competing Interest Statement

The authors have declared no competing interest.

### Author Declarations

Institutional Review Board of National Institute of Neurological Disorders and Stroke gave ethical approval under protocol 12-N-0095 Institutional Review Board of Siriraj gave ethical approval under Si 501/2017 and 601/2023 Institutional Review Board under University of Pennsylvania gave ethical approval under protocol 832955 Institutional Review Board of National Human Genome Research Institute gave ethical approval under protocol 15‐HG‐0130 Institutional Review Board of Nationwide Childrens Hospital gave ethical approval under protocol IRB11-00215

