## Supplemental Methods for "A class of deep intronic *IGHMBP2* variants activate a shared cryptic splice donor, enabling correction of select variants with a single antisense oligonucleotide"

*Splice Prediction*

Splice prediction was performed with spliceAI and Pangolin. Scores were generated using the spliceAI lookup browser from Broad Institute (https://spliceailookup.broadinstitute.org/), with max distance set to 1000bp.

*iPSC engineering and culture*

iPSCs were generated from patient derived fibroblasts using the Sendai virus reprogramming kit (Cytotune) from Thermo Fisher. iPSCs were cultured in E8-Flex media on Matrigel (corning cat # CLS354234) with 10µM ROCK inhibitor (Y-27632 dihydrochloride, Tocris Cat. No. 1254) after thawing and for 24 hours after passaging. Genomic integrity was confirmed with karyotyping on all cell lines (Cell Line Genetics).

*IGHMBP2 KO cell line*

A control iPSC line was used to generate an IGHMBP2-knockout cell line. Cells were transfected with IGHMBP2-targeting gRNA and CRISPR-Cas9, followed by single-cell cloning and knockout validation. The knockout line contains an out of frame promotor deletion and no IGHMBP2 protein is expressed on western blot.

*Neuronal culturing for immunofluorescence*

Culturing for immunofluorescence staining performed as described below. On day 3, polyornithine (PLO) coated 8 well chamber slides (overnight coating at 37°C) were rinsed with 3x PBS followed by 15ug/ml laminin coating at 37°C for two hours. For spot cultures, cells were concentrated to 20k in 1µl of NMM. Slides were dried in a laminar flow hood for 1 hour. 1µl of cells were pipetted to the top right corner of each well and allowed to adhere for 2-5min. NMM was then added, already containing treatment of choice or no treatment as needed. For standard culturing, laminin coating was removed and NMM added immediately. Cells were counted and 80k cells were plated per well.

*Fibroblast Culture and ASO treatment*

Patient-derived dermal fibroblasts were cultured in high-glucose DMEM supplemented with 10% fetal bovine serum (FBS) and 1% penicillin–streptomycin. For treatment experiments, 3 × 10⁴ cells were plated per well in 24-well plates to achieve approximately 50–60% confluency. Eight hours after plating, the medium was replaced with DMEM containing 5% FBS without penicillin–streptomycin, and either custom IGHMBP2-targeting phosphorodiamidate morpholino oligomers (PMOs) or a standard control PMO (Gene Tools) were added at the indicated concentrations together with 6 µM Endo-Porter. Cells were harvested in TRIzol 48 hours after treatment.

*RT-PCR*

RNA was extracted as per methods described below. cDNA was prepared using 1µg RNA (Applied Biosystems™ High-Capacity RNA-to-cDNA™ Kit). RT-PCR was performed using primers *GAPDH* (Forward: CCTTCATTGACCTCAACTAC and Reverse: GGAAGGCCATGCCAGTGAGC) and *IGHMBP2* (Forward: CTGAAGGCCAGAAAGTGCATCC and Reverse: GGTACTGCACCGTCAGTGTCCG) with FastStart™ High Fidelity PCR System (Roche). Reaction was separated on a 2.5% agarose gel, imaged and quantified. *IGHMBP2* values were normalized to *GAPDH*.

*Protein collection*

Cells were lysed in RIPA buffer (Thermo Scientific, 89901) with phosphatase (Sigma-Aldrich, 4906845001) and protease inhibitors (Roche, 11836170001). Cells were scraped and collected into 1.5ml Eppendorf tubes. Samples were rotated at 4 degrees for 1 hour, sonicated and centrifuged at 14000xg for 20 minutes and supernatant collected. Protein was quantified by BCA assay.

*RNA extraction*

Cells were rinsed 1x in PBS, then incubated with Trizol reagent for 2 minutes. Trizol was collected, mixed with chloroform and incubated for 2 minutes, and then centrifuged 12000xg at 4°C for 15 minutes. The upper phase was mixed with 100% ethanol and extracted using Qiagen RNeasy mini kit (cat #74104). RNA purity and concentration were determined by Nanodrop and quality measured by Agilent BioAnalyzer. All samples had a RIN > 9.5.

*Mass spectrometry*

In solution samples were reduced with 5 mM dithiothreitol at room temperature for 1hr, alkylated with 10 mM iodoacetamide for 30 min in dark, and digested with trypsin (Promega) 1:10 (w/w) at 37ºC for 18hr. Tryptic digests were cleaned with an Oasis HLB 30mg plate (Waters). ~1µg of each sample was used for the liquid chromatography-tandem mass spectrometry (LC-MS/MS) experiment. Data were acquired in data-independent mode (DIA) on an Orbitrap Ascend mass spectrometer (Thermo Fisher Scientific) coupled with a Vanquish Neo HPLC (Thermo Fisher Scientific). Peptides were separated on an ES902 column (Thermo Fisher Scientific) with the mobile phase B (0.1% formic acid in LC-MS grade acetonitrile) increasing from 3 to 28% over 73 min. The composition of mobile phase A is 0.1% formic acid in LC-MS grade water. Both MS1 and MS2 scans were performed in Orbitrap. For the MS1 scan, the following parameters were used: mass range (m/z) = 380-985; resolution = 120K; the automatic gain control (AGC) = 1e6; RF lens = 55%. The parameters for MS2 scan were: mass range (m/z) = 145-1450; resolution = 15K; isolation window (m/z) = 12; window overlap (m/z) = 1; AGC = 1e5; maximum ion injection = 40 ms. The cycle time was set at 3 sec.

*RNA Immunoprecipitation*

RNA immunoprecipitation was performed using the Magna RIP kit from Millipore (cat# 17-700) and manufacturer’s protocol. Briefly, 10x10^6^ control fibroblasts were grown to 70% confluency and collected by trypsinization, washed twice in PBS, and resuspended in RIP lysis buffer containing protease and RNase inhibitors. Magnetic beads were washed in RIP buffer and complexed with 5ug of anti-IGHMBP2 antibody (Millipore, MABE162) for 30 minutes at room temperature with rotation. Labeled beads were incubated with cell lysate overnight at 4°C with rotation. Beads were then washed a total of 6 times with RIP wash buffer and resuspended in proteinase K buffer and 1% SDS at 55°C for 30 minutes with shaking. Purified RNA was collected in phenol:chloroform:isoamyl alcohol, precipitated, and resuspended in RNase free water. Libraries were sequenced by Illumina HiSeq and the enrichment ratio of IGHMBP2 bound RNA to input was calculated.

*Immunofluorescence staining*

Cells were fixed in 4% paraformaldehyde containing 4% sucrose for 10 minutes at room temperature (RT), followed by two washes with DPBS. For growth cone analysis, cells were permeabilized with ice cold methanol for 5 min followed by 0.3% TX-100 in PBS for 10 min RT, then blocked for 1 hour at RT in 10% normal goat serum (NGS) with 0.1% Triton X-100. For other staining, cells were permeabilized and blocked together as described above. Primary antibodies were incubated overnight at 4°C in 5% NGS with 0.1% Triton X-100 using the following antibodies: mouse anti-HB9 (1:200, DSHB, 81.5C10), chicken anti-βIII tubulin (1:1000, Sigma-Aldrich # AB9354), and rabbit anti beta Actin (1:1000, GTEX, GTX109639). Following three washes with 0.1% Tween, samples were incubated with Alexa Fluor-conjugated secondary antibodies (goat anti-chicken, goat anti-mouse and goat anti-rabbit; ThermoFisher) in 5% NGS in PBS 0.1% Tween for 1 hour at RT. When assessed, 2 drops of F-actin phalloidin conjugate was added to 1 mL of secondary solution (ActinRed 555 ReadyProbe, Invitrogen R37112). After three additional washes in 0.1% Tween and one PBS wash, slides were incubated with 5ug/mL DAPI solution in PBS (Thermo Fisher) for 5 minutes, washed with PBS, once with water, and then mounted with PermaFluor Mounting Medium (ThermoFisher).

*Image acquisition and analysis*

Images were acquired using a confocal microscope (Stellaris 8, Leica). Growth cone analysis was performed using QuPath (v0.5.1-x64). F-actin and βIII tubulin stains were used to manually define regions of interest (ROI) in a blinded manner. Mean β-Actin intensity values were extracted from the ROI and statistical analysis performed with GraphPad prism (v10).

*Long read RNA seq with size selection*

Three hundred nanograms of total RNA were used to synthesize cDNA using Iso-Seq Express 2.0 Kit (Pacific Biosciences, Menlo Park, CA). The cDNA was amplified using barcoded Iso-Seq Primers (Pacific Biosciences, Menlo Park, CA) to reach the required input amount for Kinnex PCR, followed by seven parallel Kinnex PCR reactions with Kinnex primers to generate DNA fragments containing orientation-specific Kinnex segmentation sequences. The PCR-amplified Kinnex cDNA fragments were size-selected using SPRIselect beads (Beckman Coulter, Indianapolis, IN) to remove fragments below 3kb, then pooled equally and treated with Kinnex enzyme, ligated to barcoded Kinnex terminal adapters to assemble cDNA segments into an array. After removal of incomplete array, the final Kinnex library was bound to SPRQ polymerase with SPRQ Polymerase Kit, then sequenced with SPRQ Sequencing Plate and Revio SMRT cell (25M) on the Revio instrument (Pacific Biosciences, Menlo Park, CA) for 24 hours.

*Targeted long read RNAseq*

210 custom probes (120bp each) tiling 1x over canonical *IGHMBP2* exons and over the intron 8 inclusion region were purchased from IDT (xGen Custom Hybrid Panel). The targeted Iso-Seq libraries were prepared using Iso-Seq®Express Captures Using IDT xGen Lockdown Probes protocol and SMRTbell prep kit 3.0. The two targeted isoseq libraries were pooled and sequenced on a PacBio Revio using SMRTbell prep kit 3.0. Samples yielded between 6.1 to 6.4 million HiFi reads, with a mean read length of 1.13 kb and mean read quality QV37.1.

*CRISPRi Screen iPSC derivation and neuronal differentiation*

A control iPSC line (WTC11, Cat no. GM25256) for the CRISPRi screen was obtained from the Coriell Institute. For neuronal differentiation, iPSCs were engineered to stably express an inducible transcription factor cassette containing either hNIL: (neurogenin-2 NGN2, islet-1 ISL1, LIM homeobox 3 LHX3) for motor neuron differentiation or NGN2 alone for cortical neuron differentiation as previously described.*(88, 89)* The hNIL transcription factor cassette was inserted into the CLYBL safe harbor locus. Cells were engineered to express the catalytically inactive dCas9 fused to a KRAB transcriptional repression domain for CRISPRi. The dCas9-KRAB was cloned into a piggyBac transposon vector (System Biosciences) and transfected at 1:5 molar ratio of transposase to transposon and clones were expanded for analysis of dCas9 expression.*(88)*

Neuron differentiation was performed as previously described.*(89)* In brief, iPSC containing the NGN2 or NIL transcription factor cassettes were expanded in E8 Flex medium on Matrigel. For differentiation, cells were re-plated with accutase onto polyethylenimine coated dishes in Neuronal Induction Medium (NIM) containing DMEM/F12, N2 (1X), Glutamax (1X), non-essential amino acids (1X), N2 supplement (1X), 2ug/mL doxycycline, and 10uM ROCK inhibitor (Tocris). 48hrs after induction, half of the media was removed and replaced with neuronal maintenance media containing 1:1 (DMEM/F12:Neurobasal media), Glutamax (1X), non-essential amino acids (1X), N2 supplement (0.5X), B27 supplement (0.5X), NT3 supplement (10ng/mL), BDNF (10ng/mL), and Laminin (1ug/mL). Half of the media was replaced with fresh NMM and supplements as above every three days. All dishes were coated with 0.1% polyethylenimine (PEI, Sigma P3143) overnight in a 37°C incubator and subsequently washed three times with water and dried for one hour at room temperature in a laminar flow hood.

*Lentiviral Production and transduction*

CRISPRi knockdown plasmids were generated by cloning the sgRNAs into pU6-sgRNA EF1Alpha-BSD-T2A-eGFP (IGHMBP2 or control) vector. Lentiviral particles were produced using low-passage Lenti-X HEK293T cells and plated on 6 well dishes coated with poly-L-lysine. 2.5 x10^6^ cells were transfected with Opti-MEM (Thermo Fisher Scientific),3.75 uL Lipofectamine 3000 (Thermo Fisher Scientific), 5uL P3000 Enhancer Reagent (Thermo Fisher Scientific), pAdVantage, pMD2G, psPAX2, and sgRNA constructs. After 12–16 hours, the medium was replaced with fresh DMEM supplemented with 10% FBS and 6 µL ViralBoost (Alstem) per well. Viral supernatant was collected 48 hours post-transfection and centrifuged at >10,000 × g to remove cell debris. Viral concentration was performed using Lenti-X Concentrator (Takara, 631231) overnight at a ratio of 3 parts supernatant to 1 part concentrator. Virus was collected with centrifugation at 1500 x g for 45 minutes at 4°C, aliquoted and stored at −80°C. To determine the functional titer, 10,000 iPSCs per well were plated in a 96-well plate and transduced with a serial 1:2 dilution of viral supernatant, starting at a 50:50 virus supernatant:iPSC medium ratio and ten serial dilutions. Medium was replaced the next morning, and infection efficiency was assessed at 72 hours post-infection. Infection rates were quantified by fluorescence microscopy and optimal titers were defined as those achieving ~75–90% infection efficiency.

For iPSC transduction, cells were individualized and seeded at low density. Infection was performed either in suspension or after cell adhesion, depending on the experimental design. For the generation of CRISPRi knockdown lines, 5 × 10⁴ iPSCs per well were infected in a 6-well plate format. Medium was changed 8–24 hours post-infection to fresh iPSC medium containing Y-27632 (ROCK inhibitor). Infection efficiency was typically observable by 48 hours post-infection. Selection was initiated using puromycin (12 µg/mL) or blasticidin (15µg/mL).

*Genomic DNA isolation for CRISPRi Screen*

Genomic DNA was isolated using the Macherey Nagel NucleoSpin Blood columns (740950) following the manufacturer’s instructions. Cells were treated with Proteinase K in PBS and incubated at 56°C for 15 minutes and at room temperature for 30 minutes. After cooling to room temperature, 100% ethanol was added and vortexed before loading onto the columns. Samples were centrifuged at 4000 x g for 2 minutes and the flow through was discarded. Columns were washed with BQ2 buffer and DNA was eluted with pre-heated EB buffer. Elution was repeated a second time.
