## Supplementary figures and images for "A class of deep intronic *IGHMBP2* variants activate a shared cryptic splice donor, enabling correction of select variants with a single antisense oligonucleotide"

### Supplemental Figures

**Supplemental Figures**

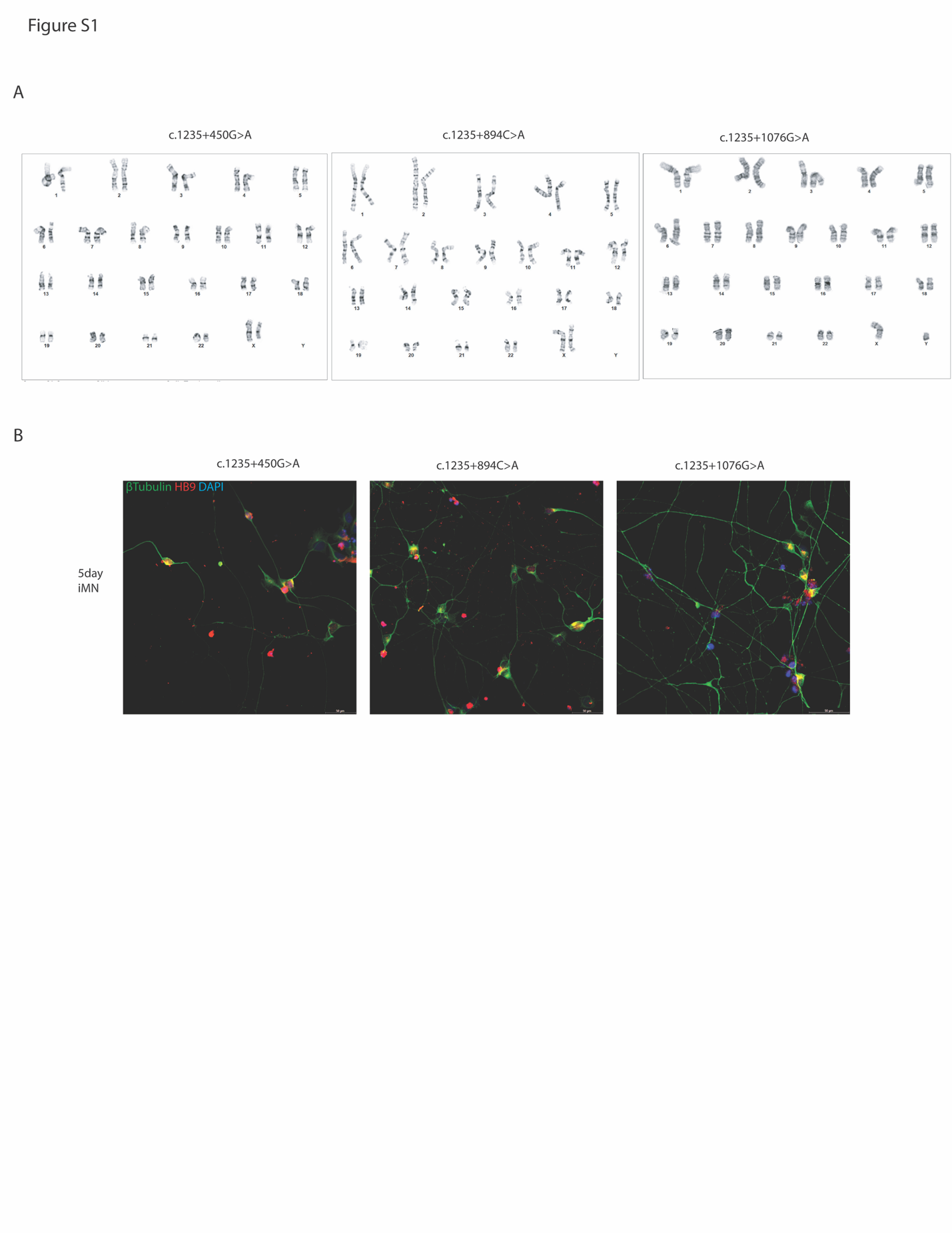

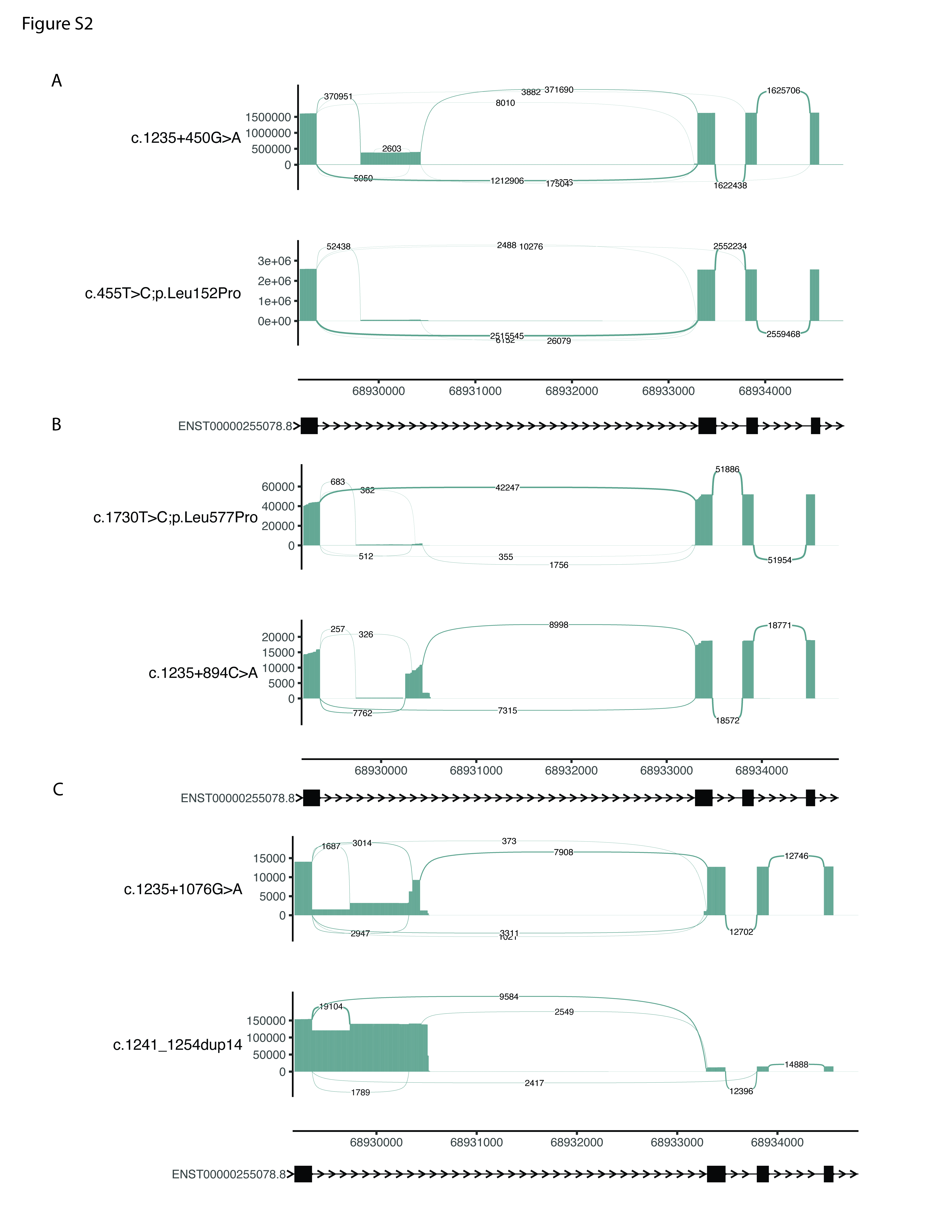

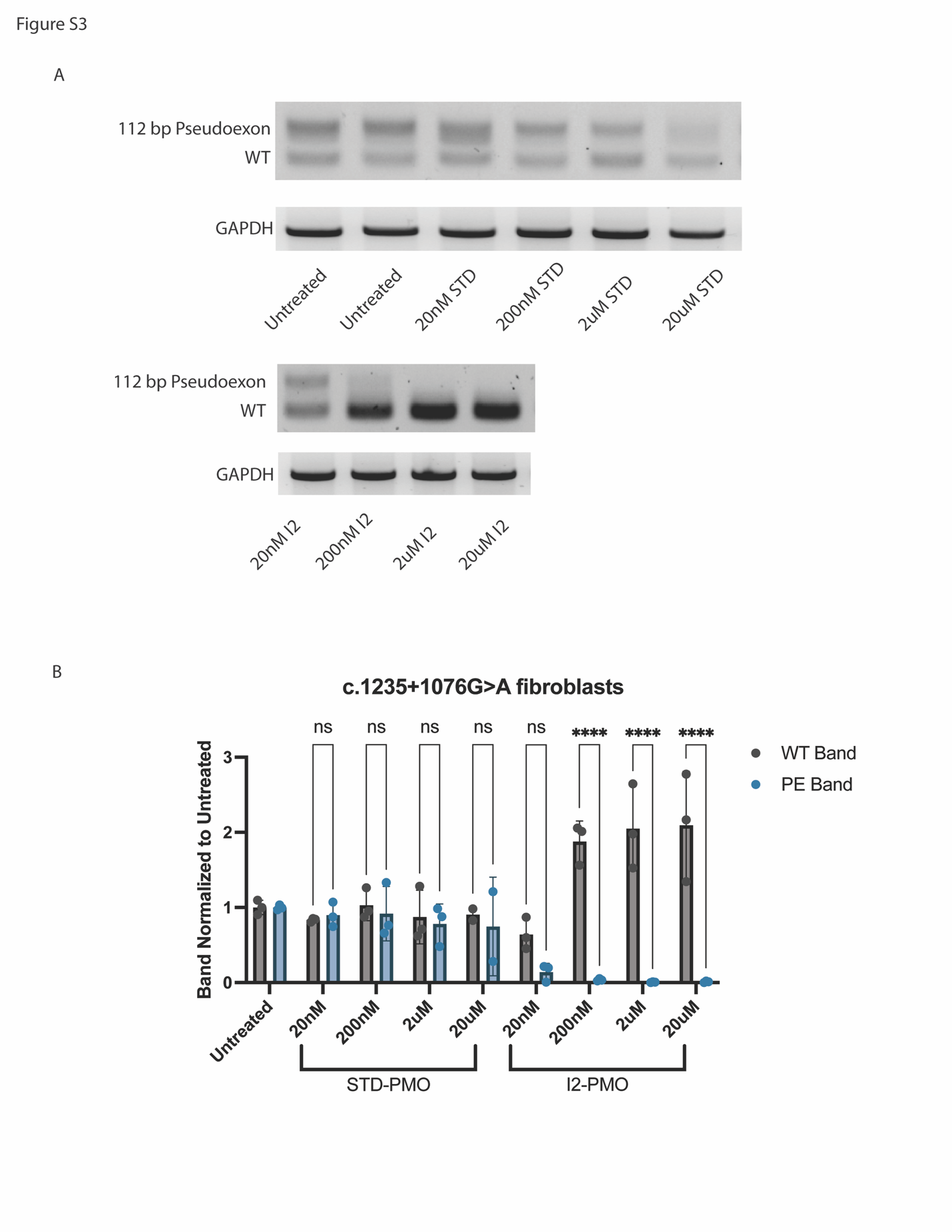

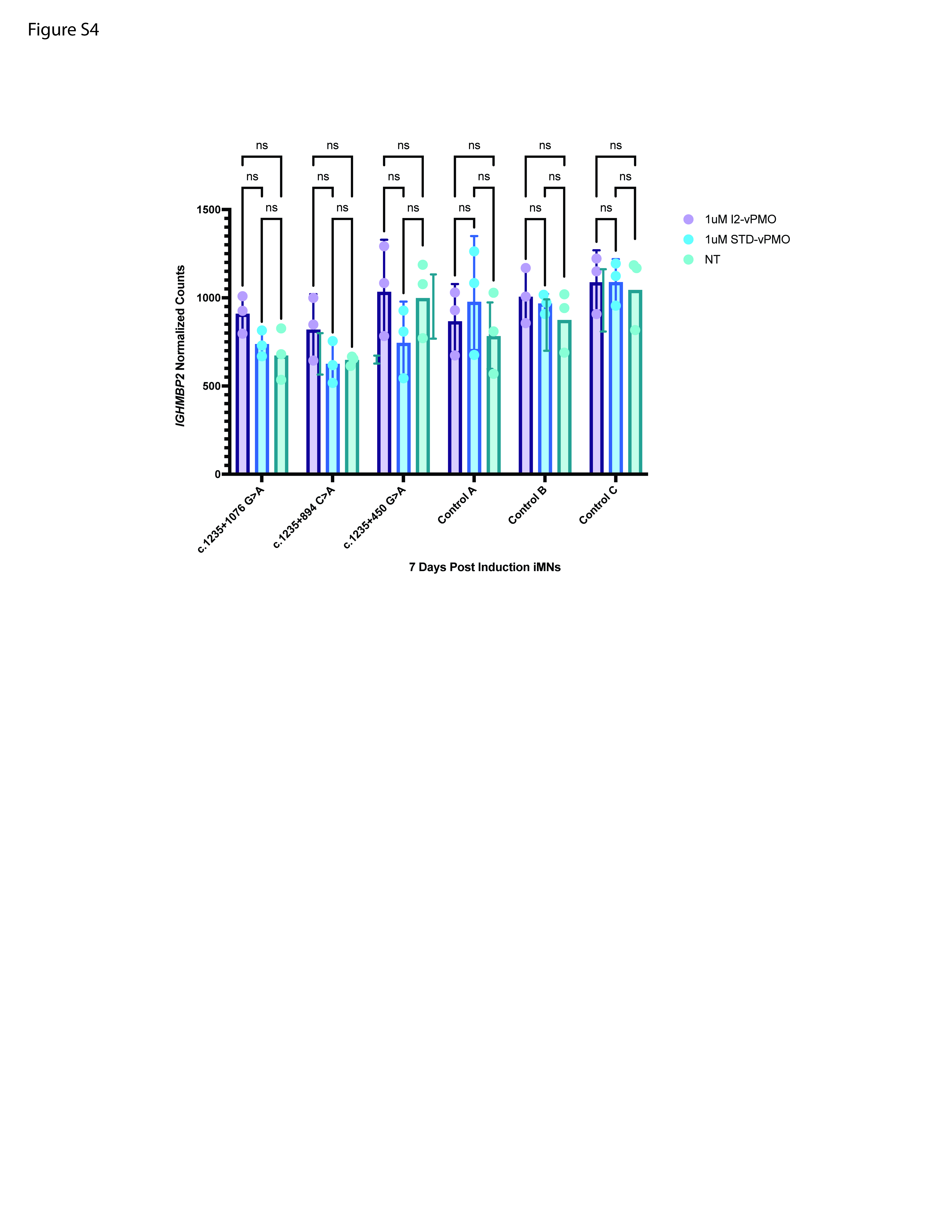

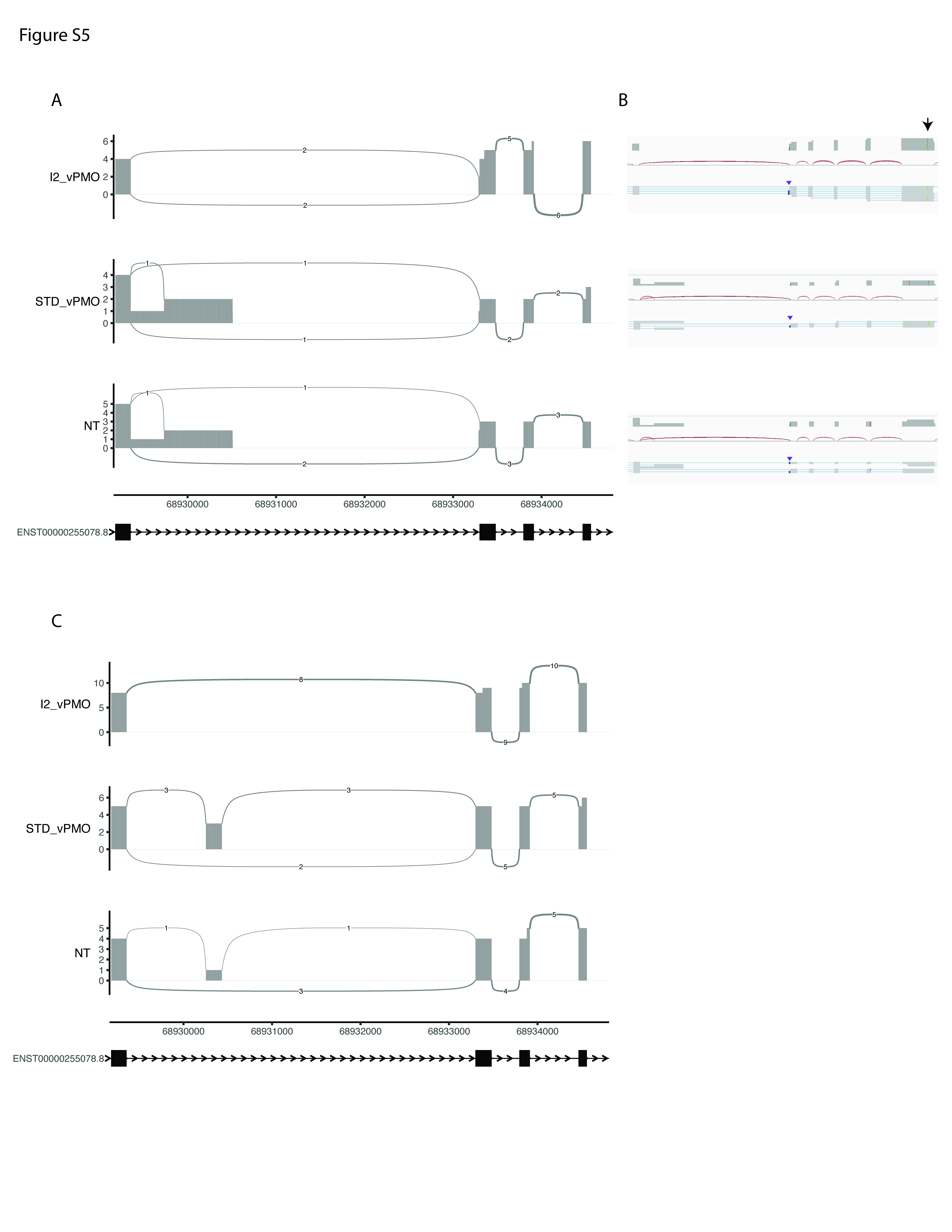

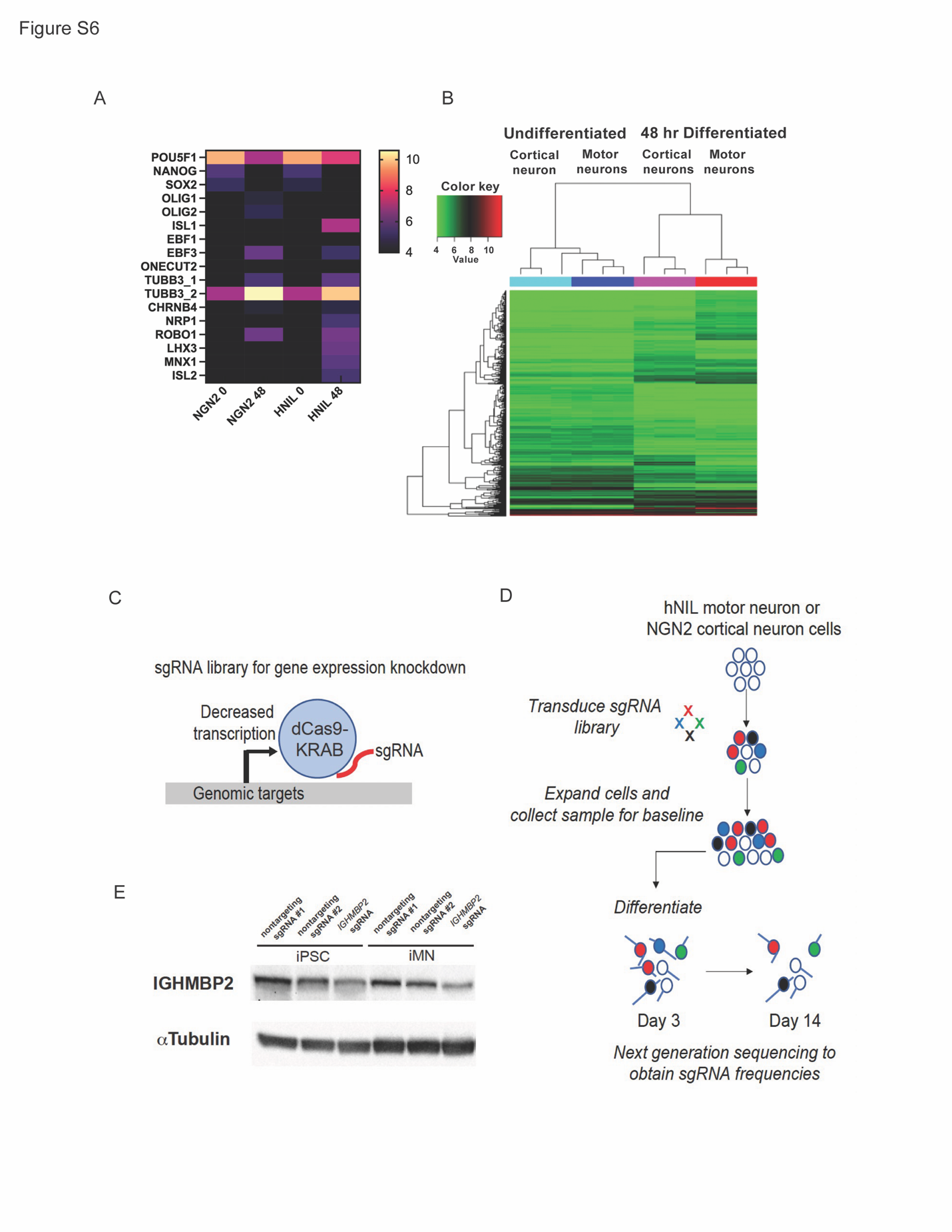

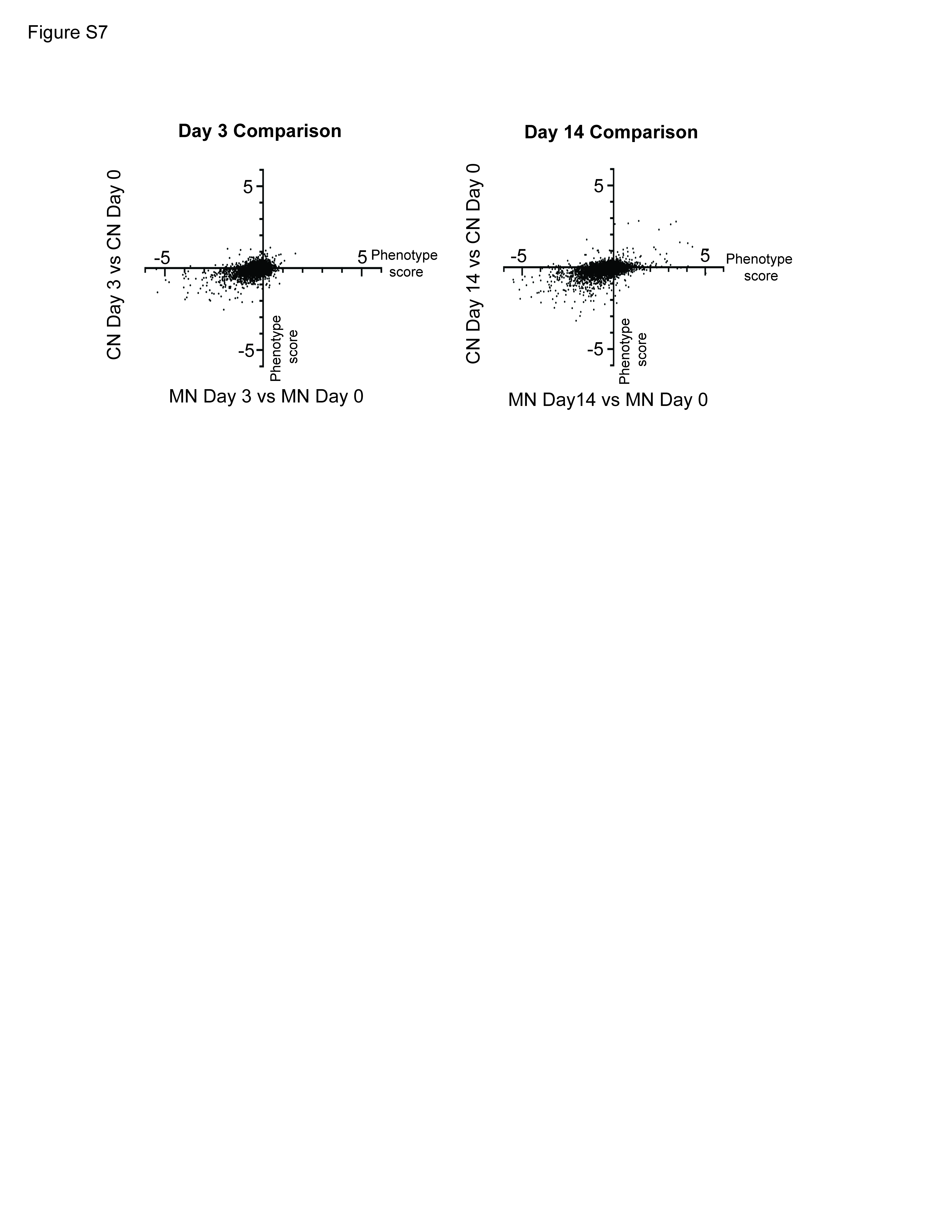

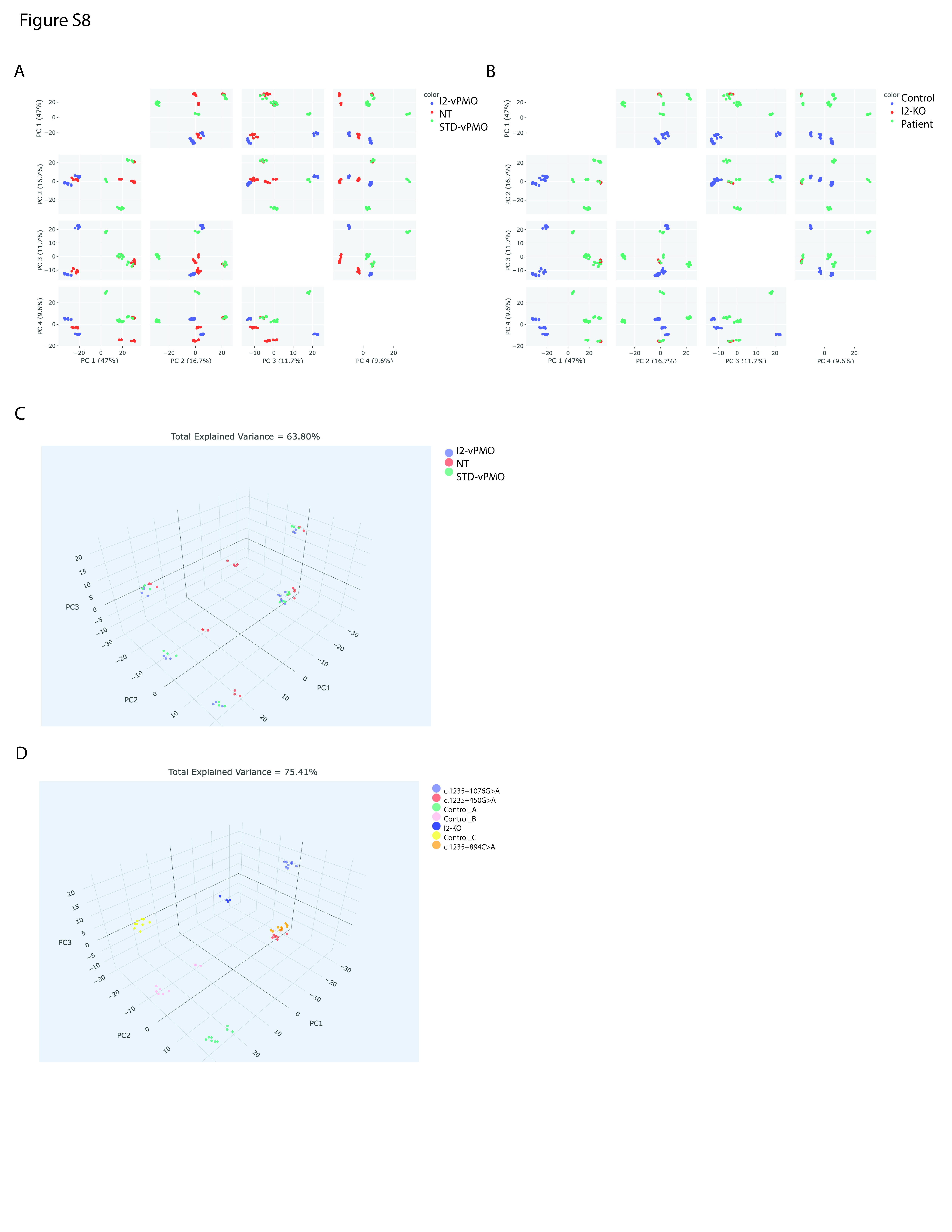

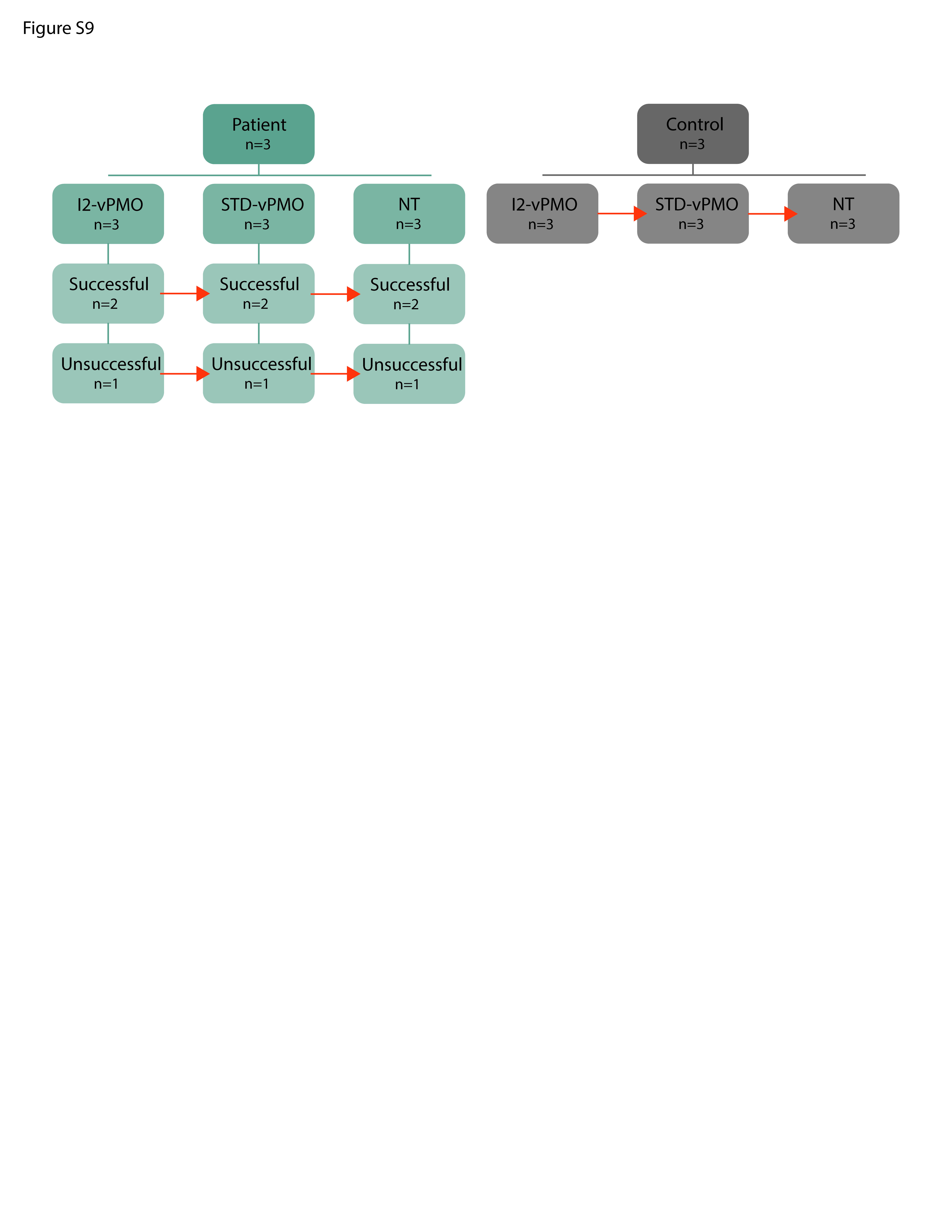

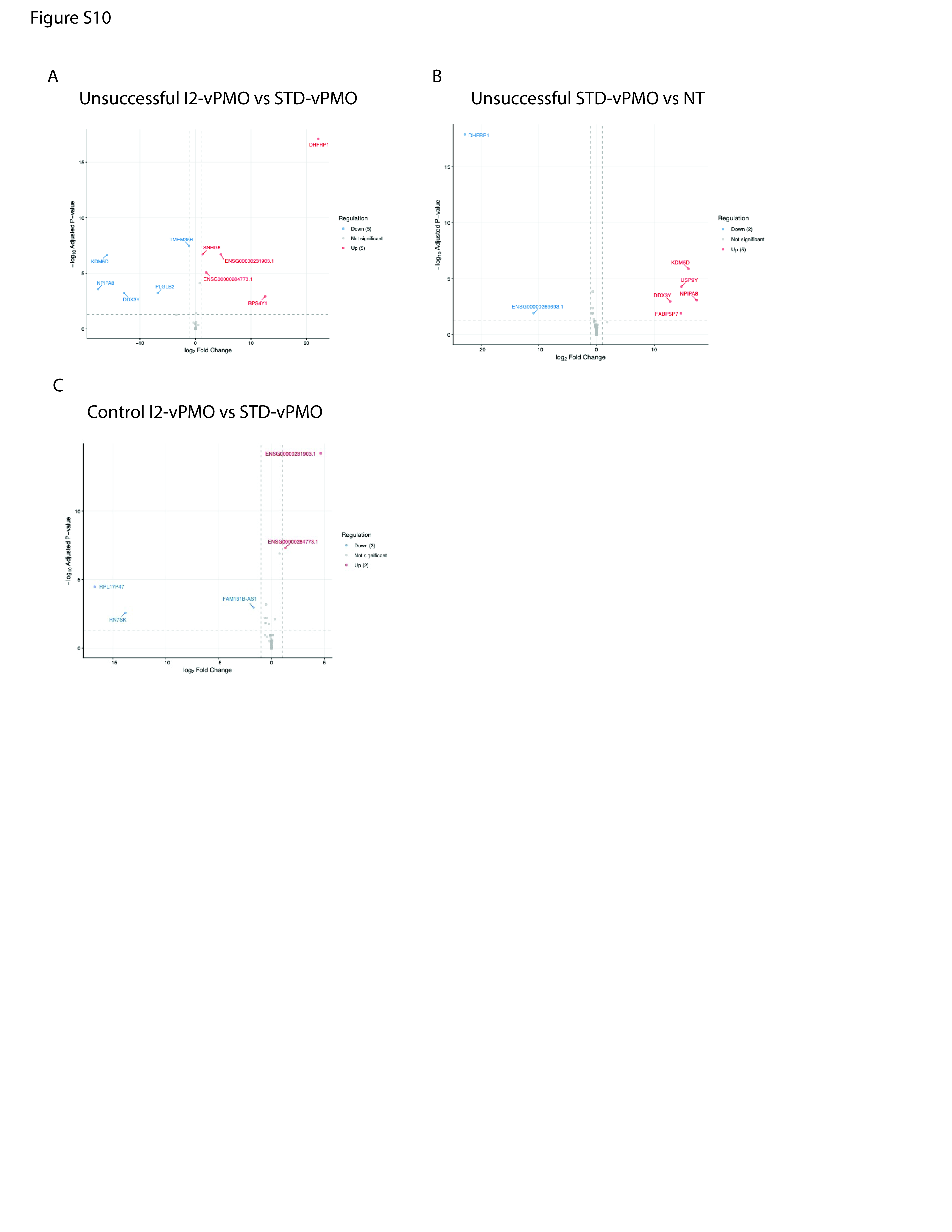

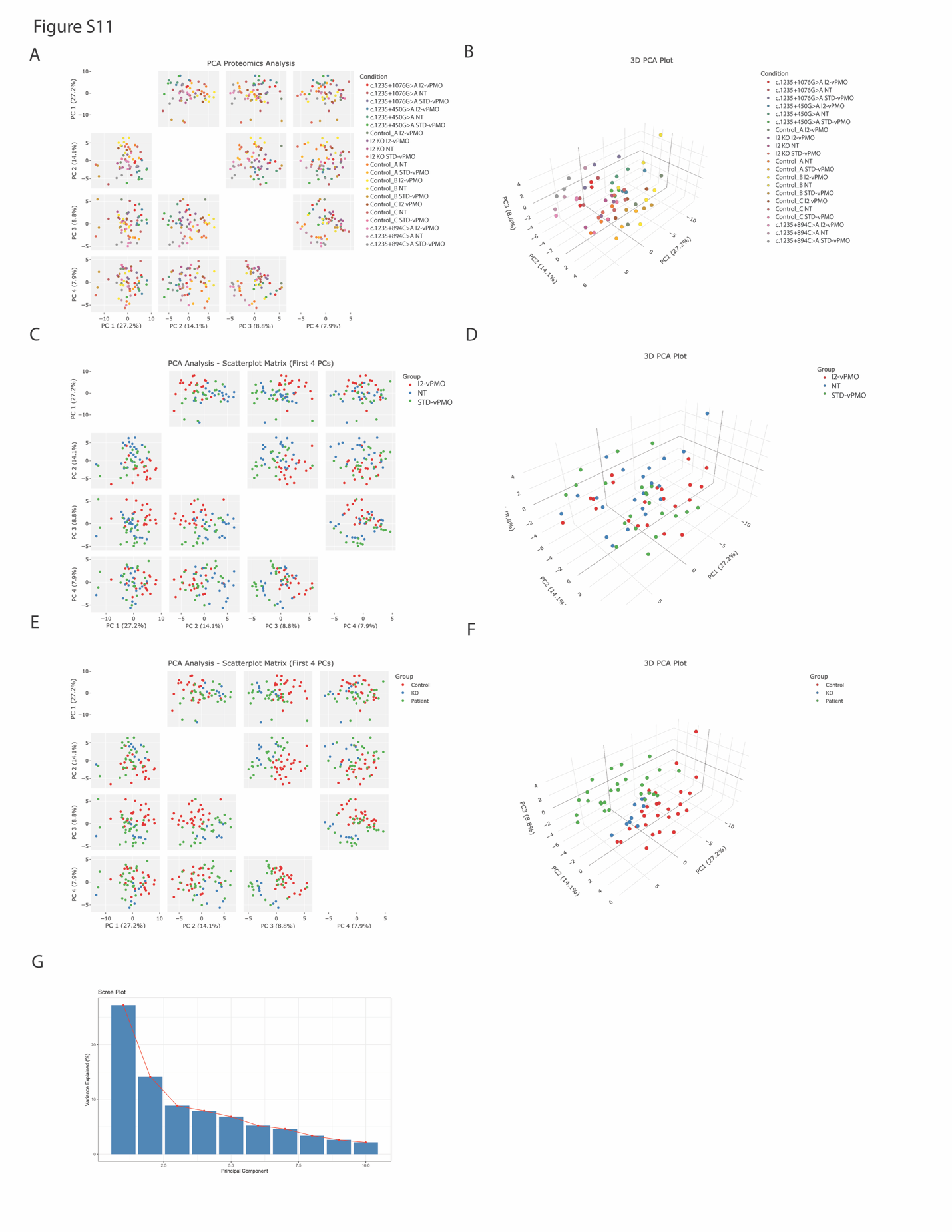

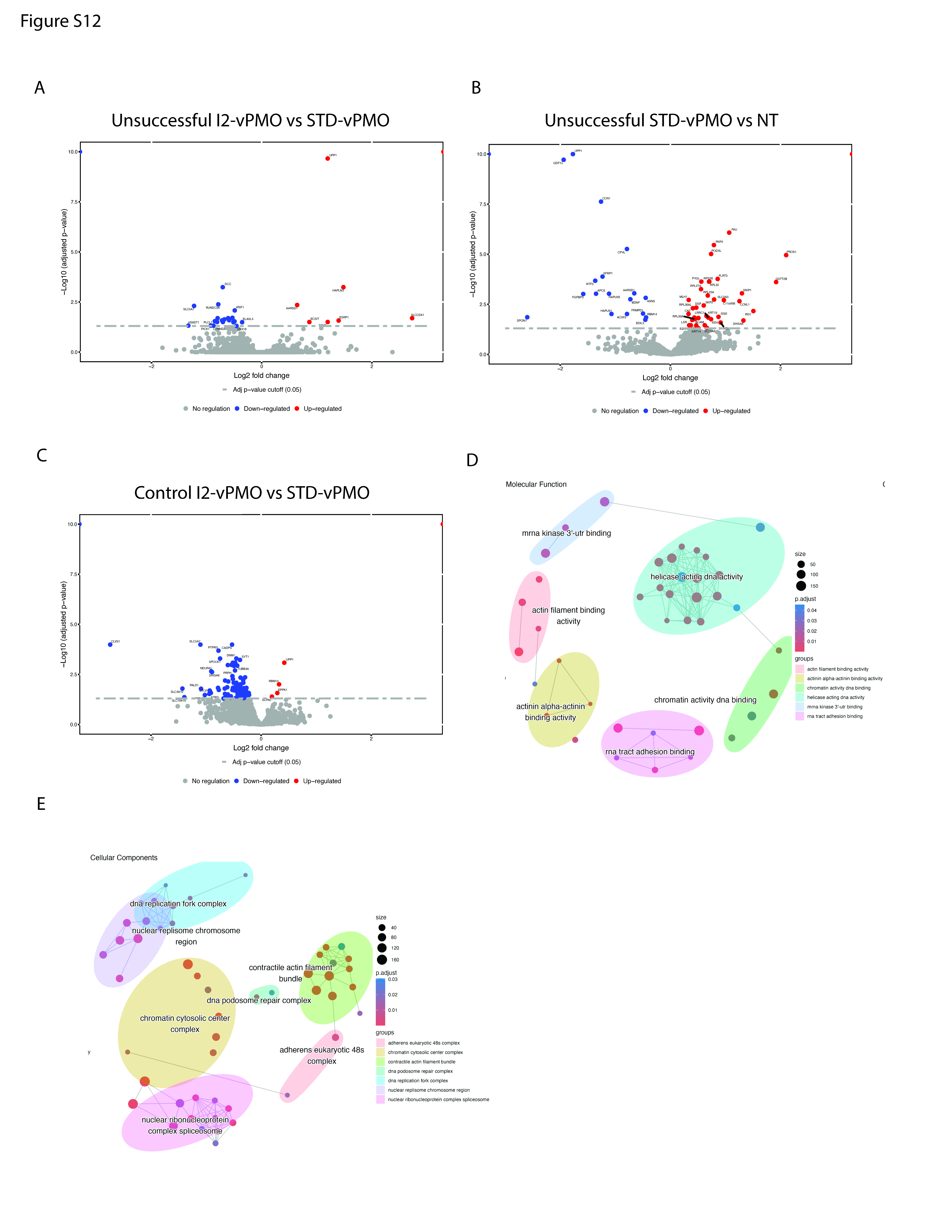

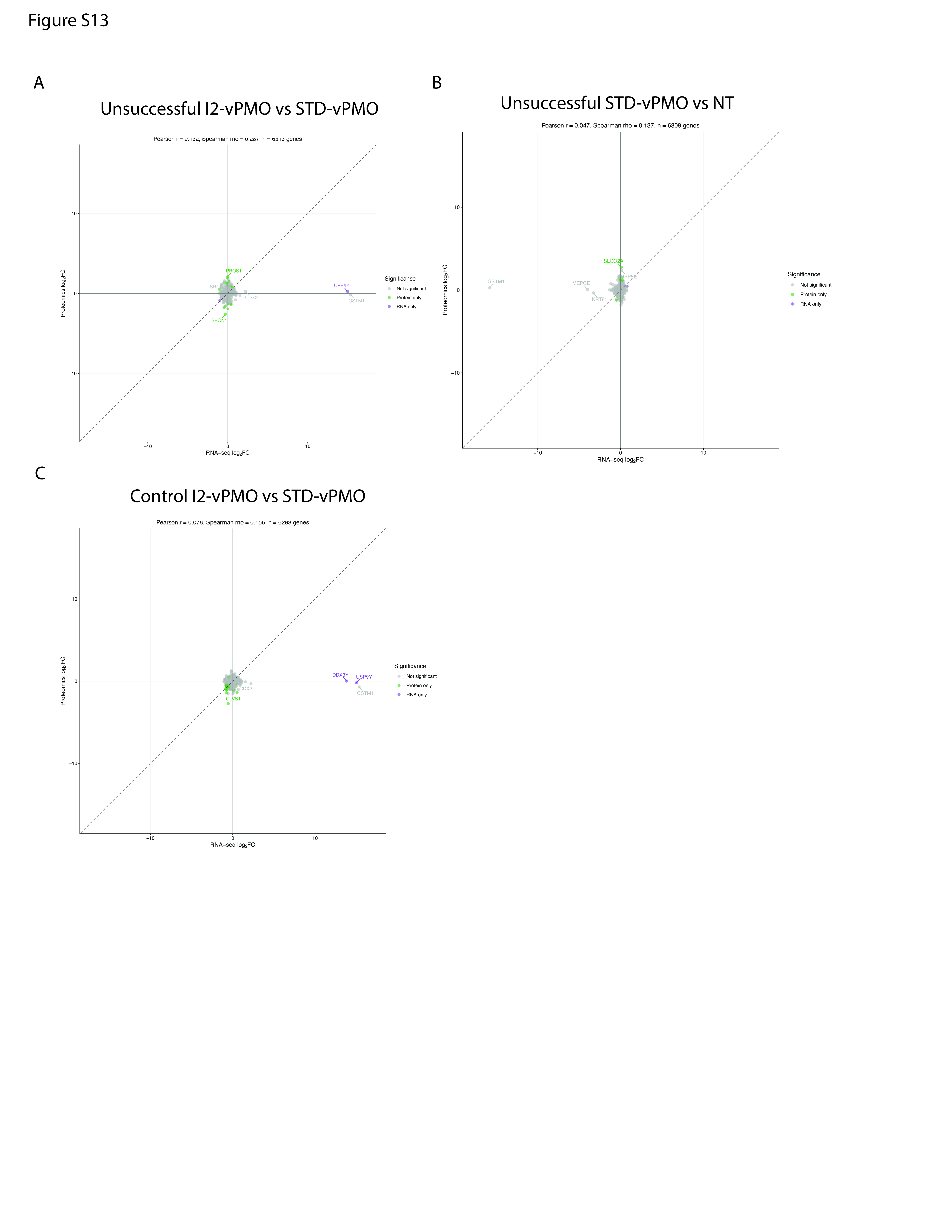

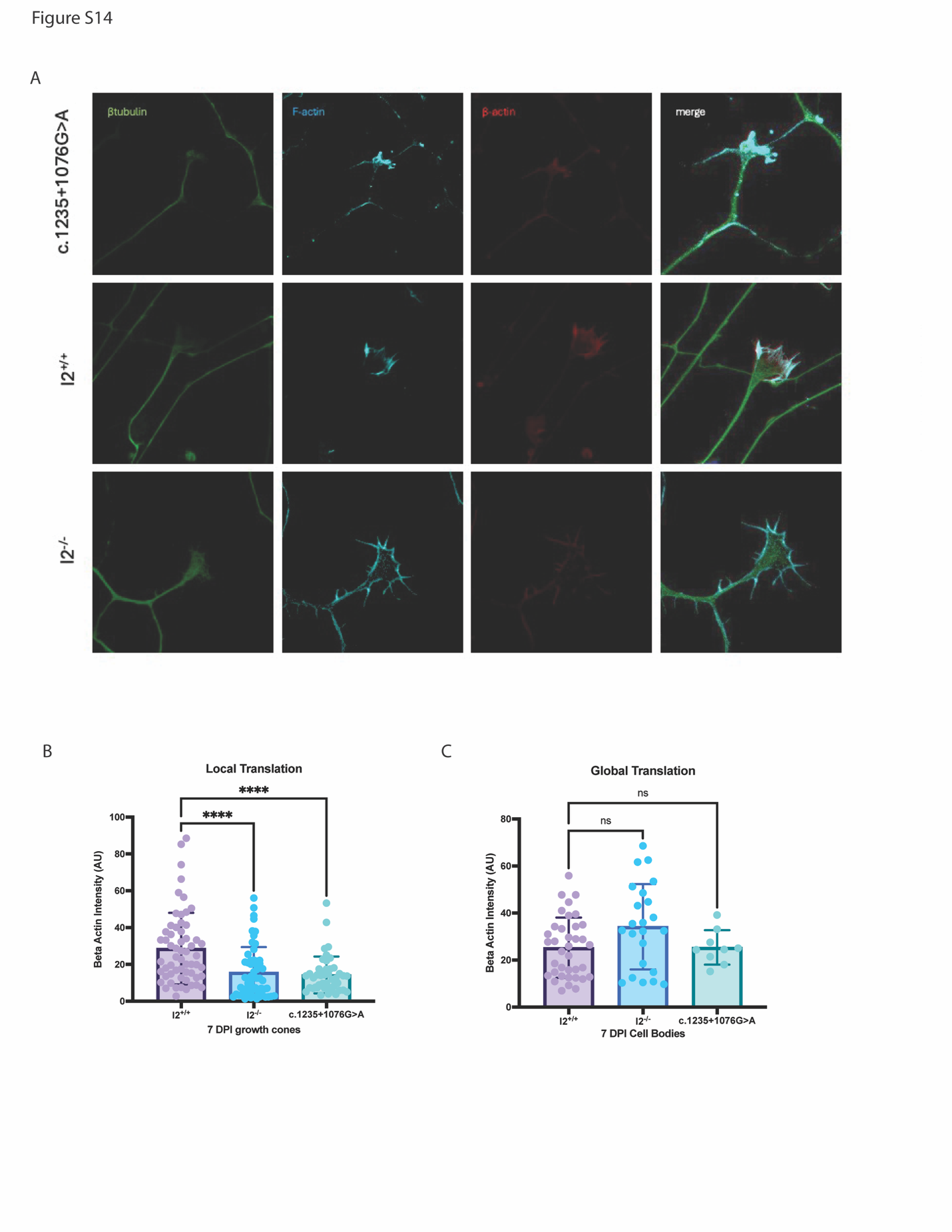
